# Multidimensional Epigenetic Clocks Reveal Physiological System-Specific Aging in Schizophrenia

**DOI:** 10.1101/2024.10.28.24316295

**Authors:** Zachary M. Harvanek, Raghav Sehgal, Daniel Borrus, Jessica Kasamoto, Ahana Priyanka, Ryan Smith, Michael J. Corley, Christiaan H. Vinkers, Marco P. Boks, Varun B. Dwaraka, Jessica Lasky-Su DSc, Albert T. Higgins-Chen

## Abstract

Schizophrenia is associated with increased age-related morbidity, mortality, and frailty, which are not entirely explained by behavioral factors. Prior studies using epigenetic clocks have suggested that schizophrenia is associated with accelerated aging, however these studies have primarily used unidimensional clocks that summarize aging as a single “biological age” score. This meta-analysis uses multidimensional epigenetic clocks that split aging into multiple scores to analyze biological aging in schizophrenia. In a meta-analysis of 7 studies with a total sample size of 1,891 patients with schizophrenia and 1,881 controls. we analyzed multidimensional epigenetic clocks, including causality-enriched CausAge clocks, physiological system-specific SystemsAge clocks, RetroelementAge, DNAmEMRAge, and multi omics-informed OMICmAge. Overall SystemsAge, DNAmEMRAge, RetroelementAge, and OMICmAge scores demonstrated increased epigenetic aging in patients with schizophrenia after strict multiple-comparison testing. Ten of the eleven SystemsAge sub-clocks corresponding to different physiological systems demonstrated increased aging, with strongest effects for Heart and Lung systems. OMICmAge DNAm proxies indicated changes in clinical biomarkers as well as novel proteins and metabolites not previously linked to schizophrenia. Most clocks demonstrated age acceleration at the first psychotic episode. Notably, clozapine use was associated with increased Heart and Inflammation aging, which may partially be driven by smoking. Most results survived Bonferroni multiple testing correction. These are the first analyses of novel multidimensional clocks in patients with schizophrenia and provide a nuanced view of aging that identifies multiple organ systems at high risk for disease in schizophrenia-related disorders.

**Key Points:** *Question:* Do novel, multidimensional epigenetic clocks demonstrate accelerated aging in schizophrenia?

*Findings:* In this meta-analysis, patients with schizophrenia-spectrum disorders demonstrated evidence of broadly accelerated aging in multiple types of epigenetic clocks. This age acceleration is particularly evident in the Heart and Lung systems and is already evident by the time of the first psychotic episode.

*Meaning:* Novel epigenetic clocks may help identify patients with schizophrenia-spectrum disorders at risk for multiple other health comorbidities.

## Introduction

Schizophrenia is a severe psychiatric disorder associated with major reductions in life expectancy of 10-20 years^1^. Much of the increased mortality stems from natural causes^2,3^ spanning multiple organ systems^4^ and multiple physical comorbidities^5-8^. Some treatments, especially clozapine, may decrease mortality from both non-natural and natural causes^9^. Increased morbidity and mortality in patients with schizophrenia is associated with accelerated aging, including elevated biomarkers of inflammation, oxidative stress, and age-associated proteins that predict mortality^10-12^.

Epigenetic clocks are commonly utilized biomarkers that estimate biological age using DNA methylation data^13-15^. Studies of schizophrenia using epigenetic clocks have shown mixed results depending on the specific clock used^15,16^. Early studies utilizing Horvath’s multi-tissue clock found no changes in epigenetic age in schizophrenia,^14,17^ likely because the clock’s training included schizophrenia samples and thus the clock is trained to ignore CpGs with altered aging patterns in schizophrenia^16^. More recent work has utilized second- and third-generation epigenetic clocks (GrimAge, PhenoAge, and DunedinPACE), which show increased epigenetic aging in schizophrenia^13,18^. These epigenetic clocks better align with epidemiologic data, are more likely to capture causal mechanisms in the aging process, and are less likely to reflect confounding cohort or period effects^19-22^.

However, traditional epigenetic clocks generally summarize aging as a single number, treating aging as unidimensional. This is problematic for studies of aging and schizophrenia, which are both complex, multi-faceted, heterogeneous phenomena in that they involve many different biological processes. Traditional clocks do not allow more nuanced questions to be asked about aging in schizophrenia. For example, could a subset of epigenetic changes seen in patients with schizophrenia actually reflect adaptive mechanisms that protect individuals with age-related disease? Do some aging processes constitute specific responses to behavioral or environmental stressors? Is the aging process uniformly accelerated in the entire body, or are specific physiologic systems disproportionately affected?

To capture multidimensionality in aging, novel epigenetic clocks have recently been developed that report a panel of biological age scores. SystemsAge is composed of 11 systems-based epigenetic clocks, developed by relating blood DNA methylation to 133 clinical and functional biomarkers organized by physiological system and then training mortality predictors for each system. Causality-enriched epigenetic clocks (CausAge) were developed using Mendelian Randomization techniques that identify CpG sites that are potentially causal for age-related traits^23^, including both detrimental (DamAge) and beneficial (AdaptAge) changes. OMICmAge was trained to predict mortality using DNA methylation data integrating 40 epigenetic biomarker proxies of proteins, metabolites, and clinical biomarkers that provide insight into specific biological processes^24^. Other interesting clocks include IntrinClock^25^, intended to capture aging unrelated to changes in cell composition, and RetroelementAge^26^, intended to capture aging at CpGs linked to retroelements.

Here, we systematically investigate these new multidimensional epigenetic clocks in 7 datasets of patients with schizophrenia and non-psychiatric controls. We hypothesized that due to increased risk of numerous diseases in schizophrenia^4,8^, most physiological systems would demonstrate increased epigenetic aging, but that specific systems would be most altered to reflect particularly high disease risks in schizophrenia (e.g. Lung for pneumonia, COPD, and smoking, or Brain for dementia risk). We use meta-analyses to assess for schizophrenia disease effects across studies, then further assessed for effects of first-episode psychosis, interactions with age, sex and smoking status, and clozapine use.

## Methods

### Selection of Datasets

We selected available epigenetic datasets of patients with schizophrenia-spectrum disorders and non-psychiatric controls^27,28^. Most datasets utilized the Illumina Infinium 450K BeadChip, though one dataset utilized the Illumina Methylation EPICv1 BeadChip. Individuals for whom chronologic age was not available were excluded from the analyses.

### Clock Calculation

Details for clock calculations can be found in Supplementary Methods. All clocks used in these analyses can be found in Supplementary Table 1. Briefly, clocks were calculated using the methylCIPHER package in R^29^ as described by^23,30,31^ or using code provided by the authors^26,32^.

### Cell-type composition estimates

Cell-type composition estimates were obtained using EpiDISH^33^. When accounting for cell-type composition, Neutrophils were dropped to avoid overfitting, and proportions of NK cells, B cells, CD4T cells, CD8T cells, Monocytes, and Eosinophils were included.

### Epigenetic smoking estimates

As smoking data was not available for most datasets, we used the GrimAge component DNAmPACKYRS, which is a proxy of smoking pack-years predicts mortality better than self-reported pack-years^20^.

### Statistical analyses

To calculate standardized effect sizes for the effect of schizophrenia, epigenetic ages were first regressed onto age, sex, and any other included covariates for the analyses (e.g., cell-type composition, smoking). These residuals were then scaled such that for the controls, the standard deviation for a given study = 1 and residual mean = 0, and then the final model was a multivariable regression of disease status, age, sex, and any other covariates regressed onto the scaled residuals. To evaluate standardized effect sizes for age-by-disease interaction, the same procedure was used. When examining clozapine effects, a similar procedure was used, limiting analyses to only individuals with schizophrenia and data regarding whether they had taken clozapine or not.

Statistical analyses were performed using R version 4.0.2^34^. Meta-analyses were performed using a random-effects model and restricted maximum likelihood estimation in the Metafor package^35^. Weights for studies were calculated as 1/SE^2^. Heatmap and scatter plots were created using ggplot2^36^. Statistics represent nominal values for consistency. However, correction for multiple comparisons via the Bonferroni method adjusts for 19 comparisons to account for all novel clocks examined (the systems-based clocks, the causality-enriched clocks, Retroelement Age, OMICmAge, IntrinClock, and DNAmEMRAge; nominal, unadjusted p of 0.00263 = adjusted p of 0.05). Similar adjustments were applied to earlier generation clocks (accounting for 14 clocks, nominal unadjusted p of 0.00357 = adjusted p of 0.05) and DNAm proxies (accounting for 40 DNAm proxies, nominal unadjusted p of 0.00125 = adjusted p of 0.05) when assessing those findings.

## Results

We identified seven different cohorts of patients (Table 1) composed of 2,210 patients with schizophrenia-spectrum disorders and 1,936 non-psychiatric controls. 549 individuals were excluded due to missing age data. Age distributions for each cohort are in Supplementary Figure 1. Two cohorts (GEO152026 and GEO152027) include individuals with first-episode psychosis (n = 716). Three cohorts (GEO116379, GEO80417, and GEO84727) include information regarding clozapine treatment (number on clozapine = 225, number confirmed not on clozapine = 406).

**Table 1:** Characteristics of included Datasets. Summary of the seven datasets analyzed here. Age indicates mean +/- SD, range in parentheses. SCZ: Schizophrenia-spectrum disorders. FEP: First-Episode Psychosis. IoPPN: Institute of Psychiatry, Psychology, and Neuroscience

| Dataset | Origin | Array Type | Controls | SCZ Cases | FEP | %Female | Age (years) | Clozapine | No Cloz |
| --- | --- | --- | --- | --- | --- | --- | --- | --- | --- |
| GEO116378 | Dutch Famine | 450K | 49 | 15 | N/A | 79.7% | 36.9 ± 16.3<br>(18.0 – 71.0) | N/A | N/A |
| GEO116379 | Chinese Famine | 450K | 79 | 74 | N/A | 50.3% | 47.8 ± 1.8<br>(44.5 – 51.0) | 35 | 39 |
| GEO147221 | Dublin Consortium | 450K | 331 | 348 | N/A | 29.0% | 41.7 ± 12.0<br>(17.0 – 70.9) | N/A | N/A |
| GEO152026 | FEP | EPICv1 | 519 | 409 | 409 | 45.2% | 35.2 ± 12.8<br>(18.0 – 64.0) | N/A | N/A |
| GEO152027 | IoPPN (King's College) - FEP | 450K | 194 | 278 | 278 | 37.8% | 28.8 ± 9.4<br>(13.0 – 72.0) | N/A | N/A |
| GEO80417 | University College London | 450K | 304 | 353 | N/A | 40.7% | 40.4 ± 15.0<br>(18.0 – 90.0) | 96 | 172 |
| GEO84727 | University of Aberdeen | 450K | 405 | 414 | N/A | 27.8% | 44.6 ± 12.9<br>(18.3 – 80.7) | 78 | 160 |

### Multidimensional Clocks are Altered in Patients with Schizophrenia

Meta-analyses demonstrated significantly increased epigenetic age in patients with schizophrenia in the AdaptAge causality-enriched clock, 10 out of 11 systems-based clocks, as well as total SystemsAge, DNAmEMRAge, OMICmAge, and RetroelementAge. Broadly, aging measures increased with chronological age, and differences between control and schizophrenia were evident in individual studies (see figures 1A and 1B for examples). The largest effects were seen for SystemsAge (β = 0.85, p = 1.7E-12), Heart (β = 0.87, p = 2.5E-12), and Lung (β = 0.82, p = 1.8E-6) clocks (Figure 1C-F). All results survived Bonferroni multiple testing correction. No significant effect was observed for the Hormone system clock (unadjusted p = 0.84), DamAge (p = 0.29), CausAge (p = 0.19), or IntrinClock (p = 0.15). These findings are summarized in the first column of Figure 1G, and all forest plots can be found in Supplementary Figure 2. When males and females are analyzed separately, we observe similar patterns (Pearson’s r = 0.96 between effect sizes calculated in males and females separately, Figure 2A).

**Figure 1:**
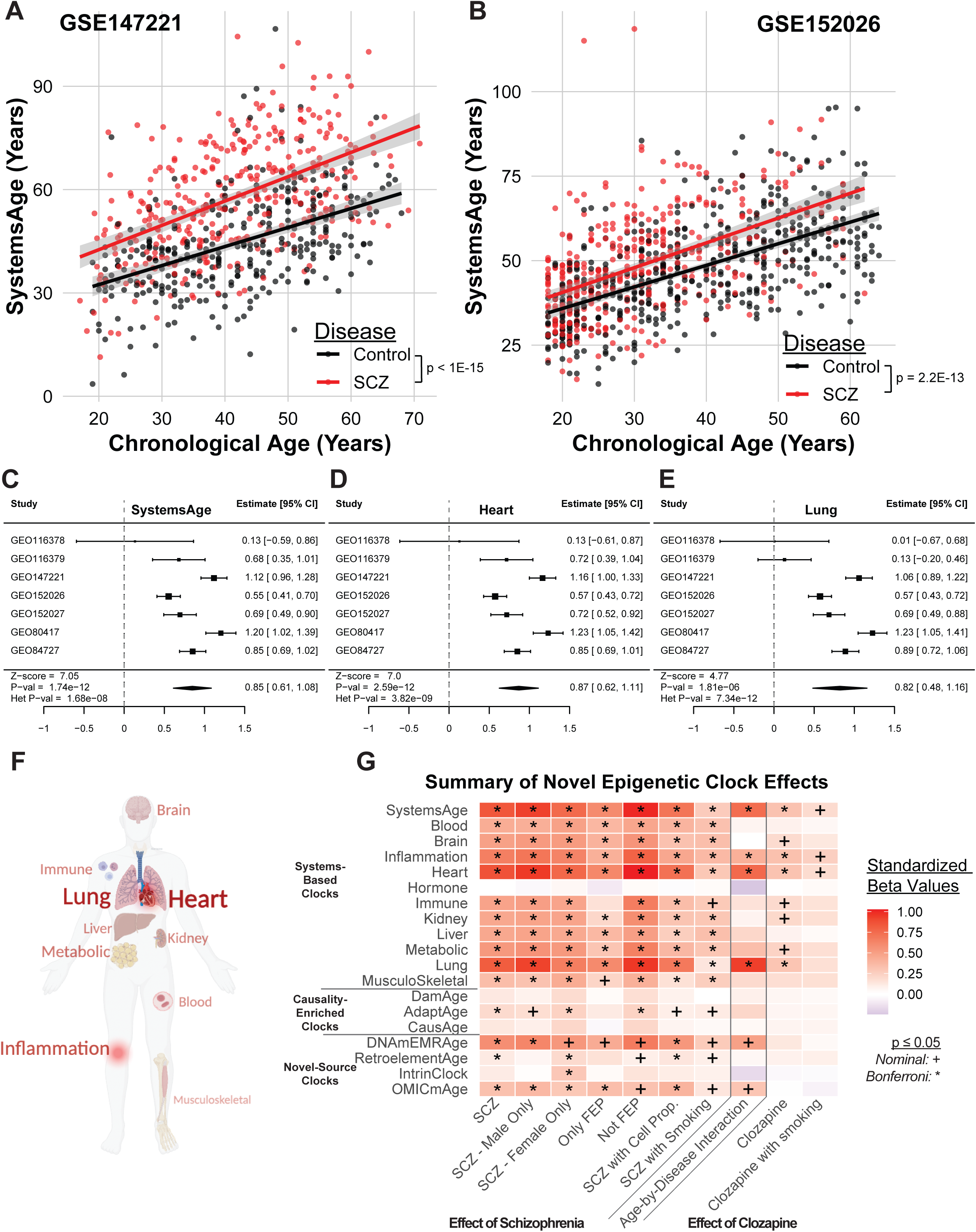
Patients with Schizophrenia have higher epigenetic age compared to controls. We observe significant age acceleration in patients with schizophrenia across multiple datasets. As examples, Total SystemsAge is plotted against chronological age for patients with Schizophrenia (red) and controls (black) for GSE147221 (A) and GSE152026 (B). Meta-analyses demonstrate this age acceleration across studies. As examples, we show Total SystemsAge (C), the Heart Clock (D), and the Lung Clock (E). Forest plots for other clocks can be found in supplementary materials. Body plot (F) demonstrates comparative effect sizes of meta-analysis in Systems-based clocks, with larger text size and bolder color indicating strongest effects. The summary heatmap (G) demonstrates standardized effect size and significance of the association between schizophrenia, schizophrenia-by-age interaction, and clozapine with all novel clocks examined. For forest plots of meta-analysis results, positive numbers indicating epigenetic age is higher in patients with schizophrenia. All p values and 95^th^ percentile confidence intervals represent nominal significance; accounting for 19 novel clocks, nominal unadjusted p of 0.00263 = Bonferroni adjusted p of 0.05.

**Figure 2:**
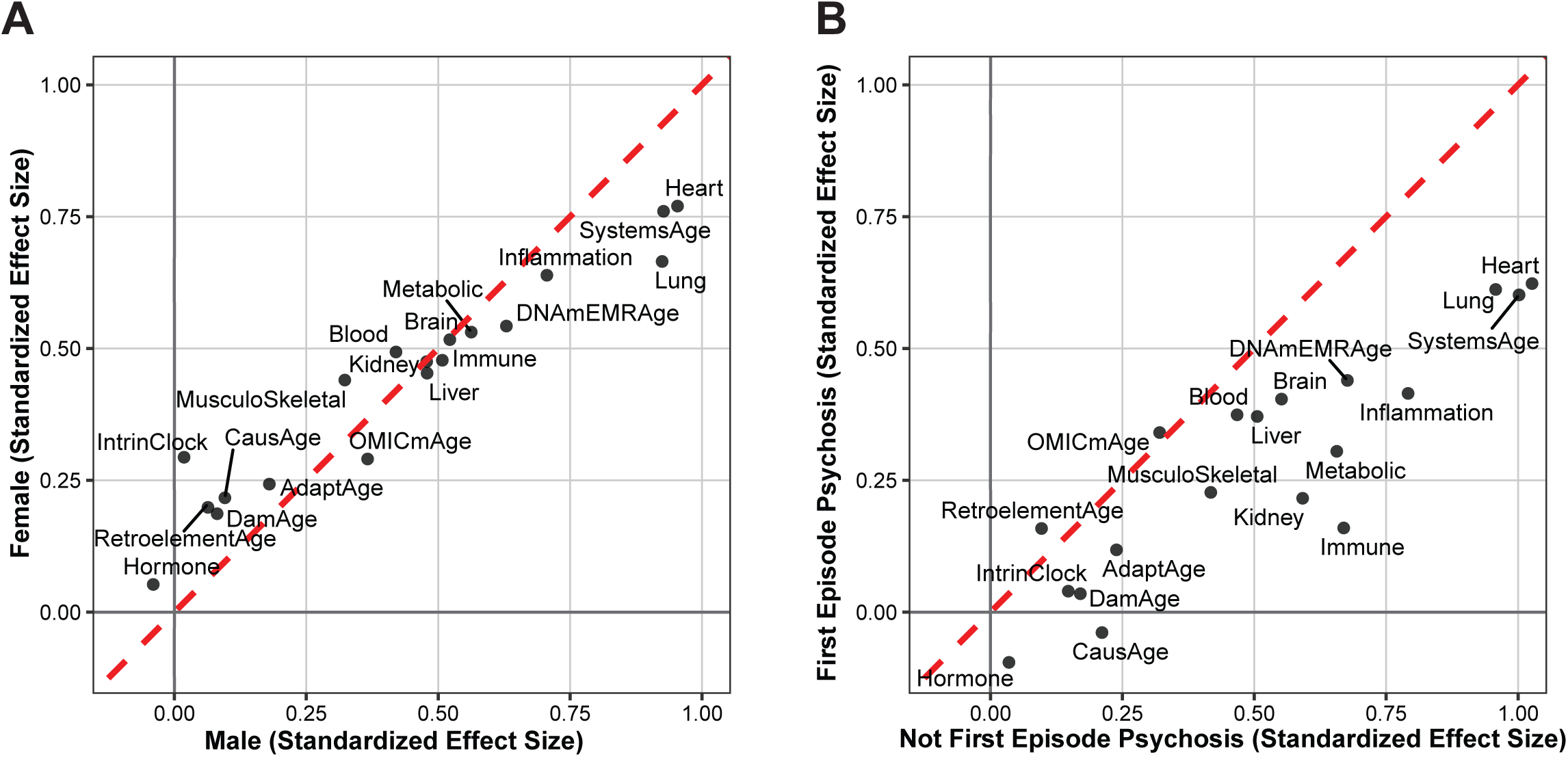
Comparison of epigenetic age effect sizes in subgroups based on sex and first episode psychosis. (A) Scatter plot demonstrates comparison of effect sizes in men versus women. (B) Scatter plot demonstrates comparison of effect sizes in patients with first-episode psychosis to non-first-episode psychosis cases. Red dashed line is the identity line (x=y).

To confirm previous findings^13,14,17,18^ using a larger meta-analysis, we repeated our analysis for unidimensional clocks. We replicated previous results demonstrating accelerated aging across most traditional clocks, except the Horvath multi-tissue and Skin and Blood clocks as expected (Supplementary Figure 3). GrimAge (both V1 and V2) showed the strongest standardized effect size of 0.98. The principal component versions of clocks^30^ largely recapitulated findings from the original clocks.

### Multidimensional Clocks are Altered in First-Episode Psychosis less than in Prevalent Schizophrenia

We asked whether these differences in epigenetic aging pre-dated psychotic symptoms by examining individuals with first-episode psychosis. None of the causality-enriched clocks demonstrate significant differences in epigenetic age in patients with first-episode psychosis compared to controls. Most of the systems-based clocks continue to show significantly higher epigenetic age at first episode psychosis except the Hormone clock, the Immune clock, and, after Bonferroni correction, the Musculoskeletal clock (Summarized in Figure 1G, “FEP” column). OMICmAge is significantly elevated in first episode psychosis, while RetroelementAge and (after Bonferroni correction) DNAmEMRAge no longer show age acceleration when just considering first-episode psychosis. First-episode psychosis is generally associated with smaller effect sizes than non-FEP, with notable exceptions of RetroelementAge and OMICmAge (Figure 2B).

We also asked whether effects of schizophrenia on clocks might change with age. Analyses excluded GEO116379 due to a narrow age range, and demonstrated a significant age-by-disease interaction in the Inflammation (p = 0.0020), Heart (p = 3.8E-5), Lung (p = 1.8E-6), and total SystemsAge (9.2E-5) clocks, and nominally significant interactions in the OMICmAge (p = 0.031), and DNAmEMRAge (p = 0.015) clocks (summarized in Figure 1G, “Age-by-Disease Interaction” column). Sensitivity analyses including GEO116379 identify significant interactions in the same clocks.

### Clozapine is associated with age acceleration across most systems-based clocks

We next sought to identify whether specific medications (i.e., Clozapine) might contribute to higher epigenetic age in schizophrenia. Patients with schizophrenia treated with clozapine showed no differences in causality-enriched clocks compared to patients with schizophrenia not treated with clozapine. However, we observed significantly higher epigenetic age in SystemsAge and 7 of 11 systems-based clocks. These effects are apparent in individual studies (See Figure 3A, B for examples) and the largest effects were observed in the total SystemsAge, Inflammation, Heart, and Lung clocks (Figure 3C-F). Smaller effects were seen for Metabolic, Immune, Kidney, and Brain that did not pass Bonferroni correction. No differences were observed in DNAmEMRAge, OMICmAge, IntrinClock, or RetroelementAge with clozapine treatment (Summarized in figure 1G, “Clozapine” column, remaining forest plots in Supplementary Figure 4).

**Figure 3:**
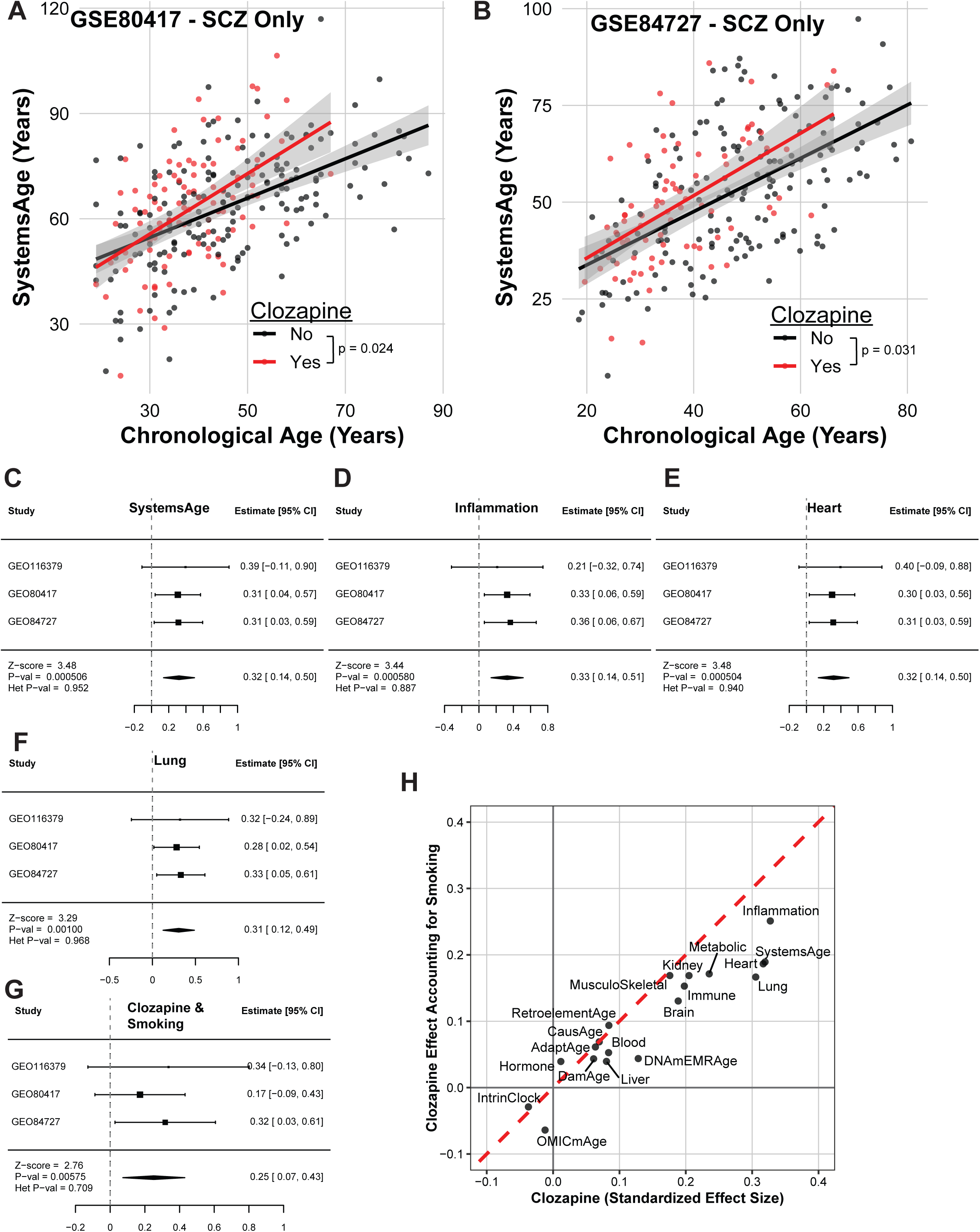
Clozapine treatment is associated with accelerated aging in the total SystemsAge, inflammation, heart, and lung clocks. Clozapine treatment is associated with age acceleration in multiple systems-based clocks. As examples, Total SystesmsAge is plotted against chronological age for patients with schizophrenia on clozapine (red) versus patients with schizophrenia not on clozapine (black) for GSE80417 (A) and GSE84727 (B). Meta-analyses demonstrate clozapine-associated age acceleration in multiple clocks, including total SystemsAge (C) and the Inflammation (D), Heart (E), and Lung (F) clocks. We do observe significantly higher values of DNAmPACKYR (an epigenetic marker of smoking) in patients on clozapine (G). (H) Scatter plot demonstrates that smoking broadly reduces effect sizes of clozapine. Red dashed line is the identity line (x=y). For forest plots of meta-analysis results, positive numbers indicating epigenetic age is higher in patients with schizophrenia on clozapine compared to patients with schizophrenia not on clozapine. All p values and 95^th^ percentile confidence intervals represent nominal significance; accounting for 19 novel clocks, nominal unadjusted p of 0.00263 = Bonferroni adjusted p of 0.05.

As we observed evidence of higher smoking rates in patients with clozapine in our study (Figure 3G), we next asked whether smoking accounted for these differences. Most systems showed weaker effects after accounting for smoking, with Lung showing the greatest attenuation as expected. After accounting for smoking, only the Inflammation, Heart, and Total SystemsAge clocks remain nominally significantly elevated with clozapine use (Figure 3H).

### Neither Cell-type composition nor Smoking account for the differences in Systems-Based Clocks in patients with Schizophrenia

We examined potential drivers of differences in epigenetic age, including cell-type composition^37^ and smoking. In general, effect sizes were lower after adjusting for cell-type composition, and AdaptAge no longer passed Bonferroni correction (p = 0.016). The conclusions from the Systems-based clocks, RetroelementAge, DNAmEMRAge, IntrinClock, and OMICmAge were unchanged after accounting for cell-type proportions (Summarized in Figure 1G, “SCZ with cell prop.” column).

When accounting for smoking, AdaptAge again shows a nominally significant association with schizophrenia (p = 0.011). Of the systems-based clocks, the Lung clock experienced the greatest decrease, with a 8.6-fold reduction of effect size, though remained significantly associated with schizophrenia (p = 0.00044). The Immune systems clock was only nominally associated with schizophrenia after correcting for smoking (p = 0.0092). All other systems remained robustly elevated in schizophrenia after correcting for smoking. RetroelementAge (p = 0.017), DNAmEMRAge (p = 0.0086), and OMICmAge (p = 0.039) showed nominal significance after accounting for smoking (Summarized in Figure 1G, “SCZ with smoking” column).

### SystemsAge Identifies Deficits in Distinct Physiologic Systems when compared to biochemical, imaging, and physical assessments

We next compared conclusions from SystemsAge to a recent study examining the health of multiple physiologic systems in neuropsychiatric disorders utilized brain imaging, physical assessment, and biochemical assays^38^. We identified little statistical correlation between these methods in terms of the effects on organ systems in schizophrenia when considering either overall estimated effect size (Figure 4A, p = 0.49, R^2^ = 0.083) or rank order (Figure 4B, p = 0.65, R^2^ = 0.036). Notably, patients with schizophrenia are more likely than controls to have increased epigenetic aging in multiple systems-based clocks (Figure 4C, p < 1E-15).

**Figure 4:**
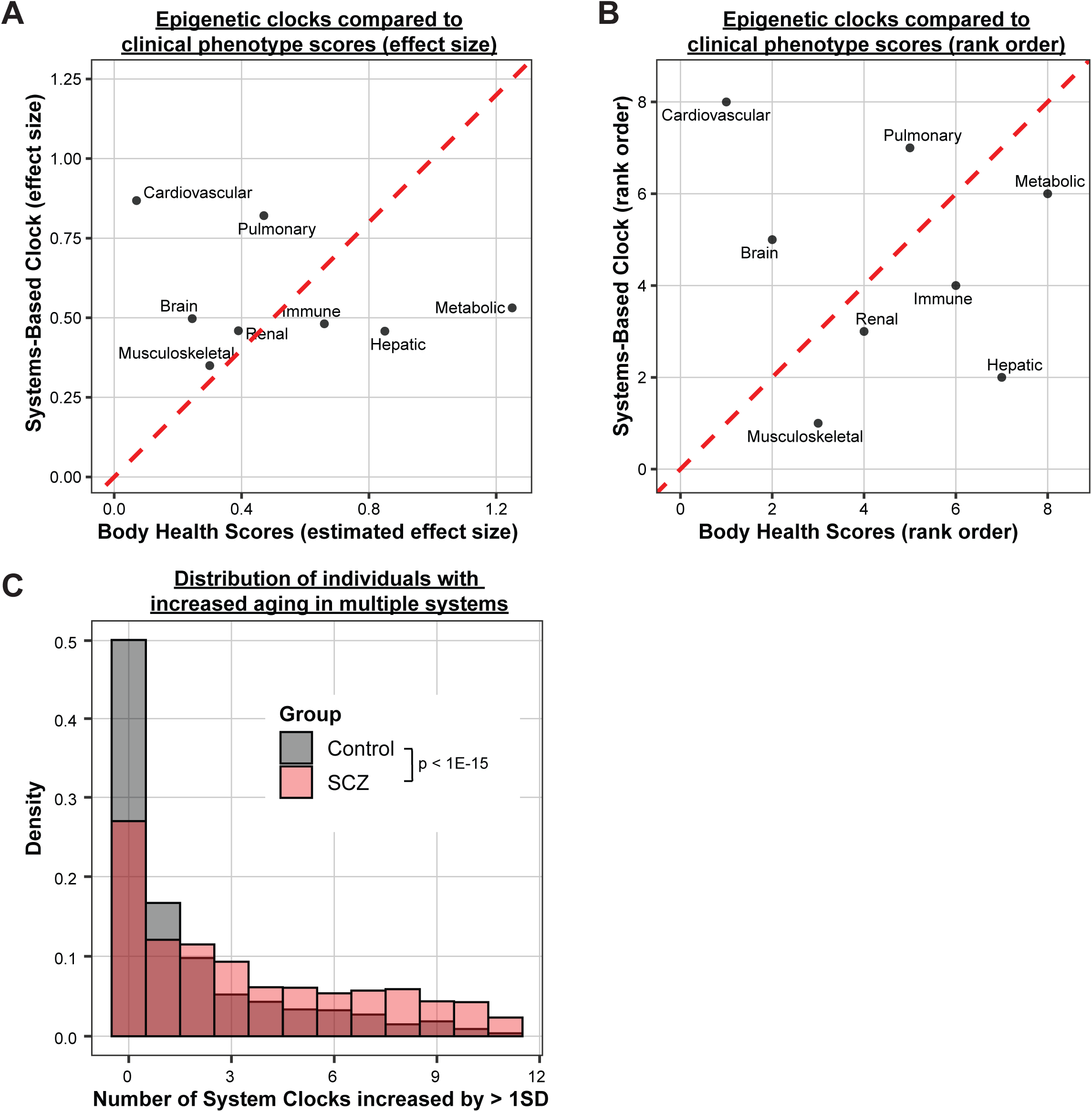
The association between epigenetic aging and schizophrenia represents a distinct phenotype from prior body health scores. (A) Scatter plot of SystemsAge standardized effect sizes versus prior reported body health scores. Red line is the identity line (x=y). (B)Scatter plot using rank-order of SystemsAge versus prior reported body health scores. Red dashed line is the identity line (x=y). (C) Histogram of # of highly accelerated systems-based clocks (age > 1SD from 0) by disease status. individuals with schizophrenia (red) or controls (black).

### Specific DNAm Proxies are altered in Schizophrenia

OMICmAge is constructed from multiple DNAm proxies for clinical biomarkers, proteins, and metabolites, which can lend potential mechanistic insight. We next asked whether these DNAm proxies showed association with schizophrenia. Of the 40 DNAm proxies examined, 19 were nominally associated with schizophrenia (nominal p ≤ 0.05), 10 of which pass Bonferroni correction (nominal p ≤ 0.00125; Figure 5A). Broadly similar patterns were observed in men and women. Remarkably, 12 DNAm proxies were significant in the first-episode psychosis datasets, 3 of which met Bonferroni: ITIH3, RNAS1, and Mimecan. In the non-first episode psychosis datasets, the majority of DNAm proxies showed at least nominal significance. Correction for cell-type proportions and smoking generally blunted effect sizes. Of the 40 DNAm proxies, 6 showed at least nominally significant associations in all models (the full sample, male/female subsets, FEP/no-FEP subsets, after adjusting for cell-type proportions, and after adjusting for smoking): Vanillactate, H2B1C, Mimecan, RNAS1, Gluconate, and ITIH3. No DNAm proxies passed Bonferroni correction for age-by-schizophrenia interactions or treatment with clozapine.

**Figure 5:**
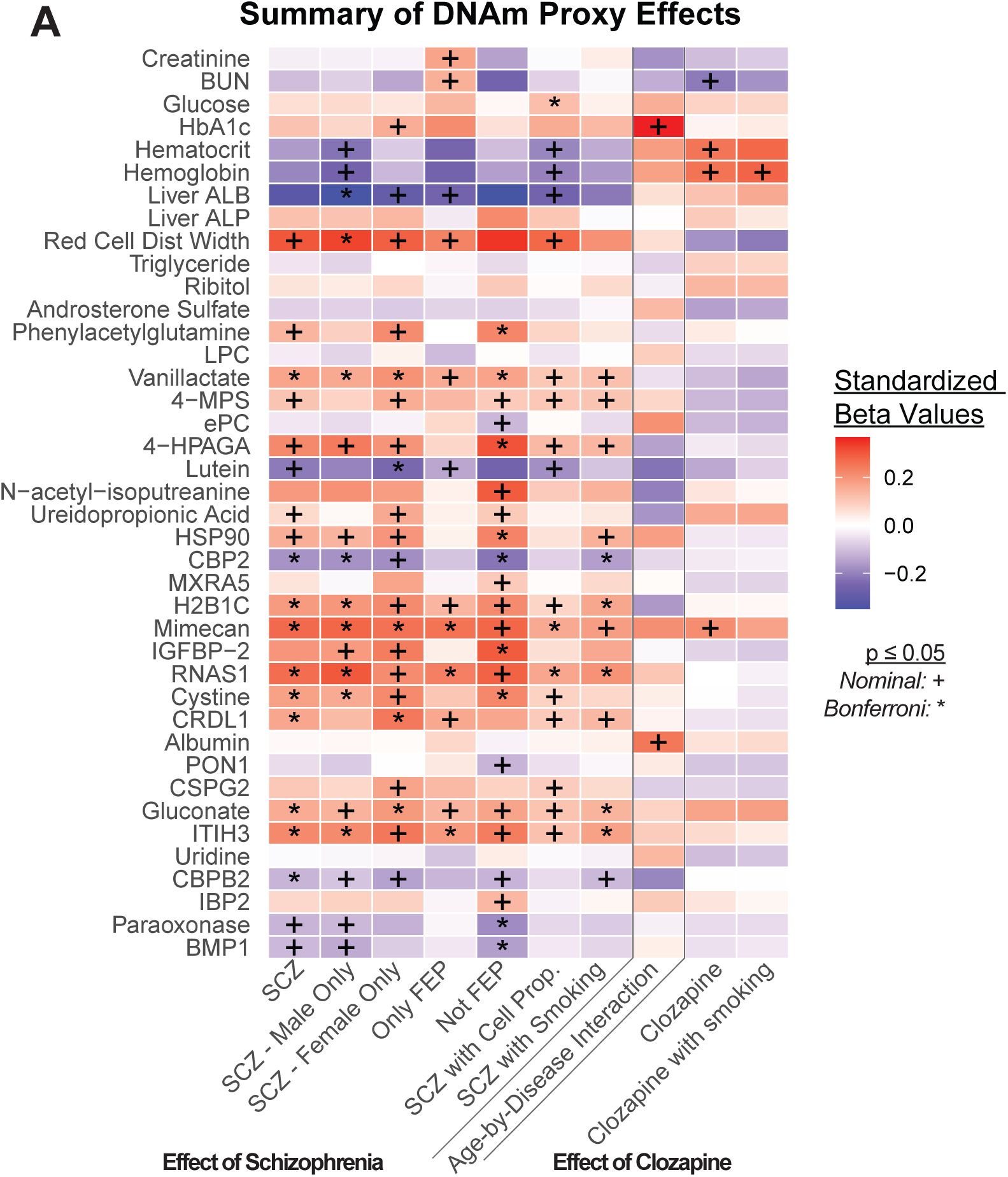
DNAm Proxies show significant associations with schizophrenia. (A) Heatmap demonstrating effect size and significance of the association between schizophrenia, schizophrenia-by-age interaction, and clozapine with DNAm Proxies from OMICmAge. Accounting for 40 DNAm proxies, nominal unadjusted p of 0.00125 = Bonferroni adjusted p of 0.05. Forest plots can be found in supplementary materials.

## Discussion

Here, we show that schizophrenia is associated with broad accelerated aging in multidimensional epigenetic clocks. These clocks provide a far more nuanced picture of aging through subscores that each capture different aspects of aging. These subscores reveal schizophrenia is characterized by adaptive age-related changes, accelerated aging in most physiological systems, many age-related metabolites and proteins, as well as retroelements. Critically, the large sample size and meta-analytic design of this study allowed us to obtain robust results that generally pass strict Bonferroni multiple testing correction even when examining numerous clocks simultaneously. Sensitivity analyses indicated the observed effects are generally independent of sex and robust correction for cell-type composition and smoking. Interestingly, smoking, clozapine, and first-episode psychosis each affected specific clock subscores, highlighting the utility of multidimensional clocks in disentangling the effects of different clinical variables on aging.

Importantly, the multidimensional clocks correlate with the known epidemiology of schizophrenia. Large studies including meta-analyses have demonstrated that individuals with schizophrenia have increased disease and mortality rates from natural causes covering every physiological system^4,8^. Accordingly, 10/11 of the SystemsAge subscores are increased in schizophrenia. Because each SystemsAge subscore has specific associations with outcomes related to that system^31^ (e.g. lung cancer for Lung, cognition for Brain, diabetes for Musculoskeletal and Metabolic), SystemsAge could help explain the increased disease and mortality risks in schizophrenia. The only score not increased in schizophrenia is Hormone, which was previously reported to be most related to thyroid disease and cancer risk^31^. Accordingly, thyroid disease is not elevated in schizophrenia^8^, and cancer shows the smallest increased risk in Correll et al^4^. The lack of age acceleration in the hormone subscore strengthens the case for the use of multi-dimensional clocks, as it suggests they maintain specificity even in diseases such as schizophrenia with significant comorbidities. As expected, patients with schizophrenia are more likely to show multi-system accelerated aging, consistent with previous results showing a 69% increased risk of multimorbidity in schizophrenia^39^. Importantly, SystemsAge can better capture the heterogeneity of risk in schizophrenia and thus could identify which diseases individuals with schizophrenia are at greatest risk for.

A recent study by Tian et al. developed system scores using clinical data and examined changes in schizophrenia^38^. Interestingly, while we found that Heart and Lung were the scores with greatest increase in schizophrenia, Tian et al. found nearly no change in their Cardiovascular score and a modest change in the Pulmonary score. The Cardiovascular score from Tian et al. utilized heart rate, blood pressure, and arterial stiffness, while the SystemsAge Heart score includes DNAm proxies of BMI, smoking, blood pressure, clinical history, and mortality-associated proteins. Clinical experience is more consistent with markedly increased heart and lung aging in schizophrenia, given the high rates of smoking^40^, known adverse effects associated with antipsychotic medications^41^ (weight gain, myocardial infarction, pneumonia), and elevated mortality risk due to pneumonia (RR 7), any respiratory cause (RR 3.75) and cardio-cerebrovascular causes (RR 3.47)^4^. Given the discrepant results from the two methods, it will be interesting to determine if combining clinical and laboratory-based risk factors with epigenetic scores may provide complementary information on risks of comorbidities in patients with schizophrenia.

Increased epigenetic age was noted at first-episode psychosis, suggesting many changes are detectable early in the disease. However, effect sizes were smaller in first-episode psychosis compared to prevalent schizophrenia for nearly all clocks. Interestingly, only a subset of clocks accelerated over time as suggested by age-by-disease interactions - these were the Heart, Lung, Inflammation, full SystemsAge, OMICmAge, and DNAmEMRAge clocks. It is possible changes in these systems reflect the cumulative effects of factors associated with psychosis (e.g., smoking, stress) and treatment of psychosis (e.g., atypical antipsychotics). This suggests that older individuals with schizophrenia may be particularly vulnerable to a cardiovascular, pulmonary, and inflammatory age-related diseases.

Clozapine has unique treatment benefits but also greater metabolic side effects and can induce serious cardiac, hematologic, and neurological adverse events. We find that clozapine treatment is associated with acceleration in SystemsAge, multiple SystemsAge subclocks especially Inflammation and Heart but also in Metabolic, Kidney, Immune, and Brain, as well as in multiple hematologic markers of OMICmAge. These changes may reflect the side effect profile of clozapine, and indeed longitudinal studies have shown that clozapine can directly impact the methylome^42^. However, there are other explanations: treatment-resistant individuals, in whom clozapine is primarily used, may represent a distinct clinical population with unique risks^43^. Notably, prior meta-analyses have suggested that clozapine is associated with decreased mortality in patients with schizophrenia, despite its known effects on cardiometabolic risk factors^4^. Further longitudinal studies are needed to determine potential causal relationships between clozapine, epigenetic aging, and mortality.

Our analysis of OMICmAge reveals novel insights into clinical biomarkers, proteins and metabolites altered in schizophrenia. Previously, we showed that DNAm proxies of serum B2M, Cystatin C, GDF-15, TIMP-1, ADM, and PAI-1 (components of GrimAge) are elevated in schizophrenia, which matches the literature concerning these proteins^13^. This suggests DNAm proxies can be used for discovery. In some cases, the DNAm proxies may even be more useful. Prior results showed stronger associations with mortality for a DNAm proxy of smoking pack-years compared to self-reported smoking pack-years^20^, and stronger associations with brain health outcomes for a DNAm proxy of CRP compared to directly measured CRP^44^. OMICmAge predicted changes in multiple clinical biomarkers which were often sex-specific. While both men and women demonstrated higher RDW and lower liver albumin, men showed associations with decreased hemoglobin and hematocrit, and women showed elevated hemoglobin A1C. These results are generally consistent with anemia, malnutrition, other comorbidities, and medication effects in schizophrenia^45,46^, although further explorations into whether the sex-specific differences in the DNAm proxies seen here translate clinically are necessary. Notably, higher RDW and lower albumin have been associated with increased mortality in the general population in both NHANES and the UK Biobank^47^. IGFBP2 was found to be increased in both men and women, (although not in the full sample), consistent with prior literature^48^. IGFBP-2 may play a role in increased metabolic risk and altered synaptic plasticity in schizophrenia or treatment^48,49^. Potentially novel proteins and metabolites that could play a role in features of schizophrenia include (4-hydroxy)phenylacetylglutamine (heart disease^50^), vanillactate (heart disease^51^), carboxypeptidase B2 (thromboembolic disease^52^), histone H2B type 1-K (cellular senescence^53^), mimecan (food intake^54^), and chordin-like protein 1 (cognitive decline^55^, adipogenesis^56^). Notably, directionality is not necessarily consistent with prior literature – for example we found increased DNAm-predicted cystine though a prior study found reduced directly measured cystine^57^, suggesting a complex relationship between cystine, DNAm and schizophrenia. These metabolites may also play roles in schizophrenia risk – we identified increased DNAm-predicted ITIH3 in all our models, including those just examining first episode psychosis. Previous genome wide studies have implicated multiple SNPs within ITIH3 in schizophrenia. While one is a missense variant, another SNP is in an intron and does not seem to affect ITIH3 expression^58,59^. The biology of these proteins and metabolites in schizophrenia represent fertile areas for future investigation.

Limitations of our study include its cross-sectional nature and limited datasets for first-episode psychosis and clozapine. The absence of patients with schizophrenia without medication prevents analyses of general anti-psychotic treatment and epigenetic aging. Future longitudinal and interventional studies with more detailed phenotypic data will be needed to determine whether antipsychotics or other factors associated with schizophrenia contribute to accelerated aging.

## Conclusion

In this meta-analysis, we identify a rich tapestry of accelerated epigenetic aging in schizophrenia-spectrum disorders. Specific physiological systems are particularly affected, changes can be either damaging or adaptive changes, and many age-related metabolites, proteins, and retroelements are affected. These findings are robust after strict multiple testing correction and correcting for covariates. Factors such as smoking, first-episode psychosis, and clozapine have effects on particular subsets of clocks. These clocks may complement clinical data in identifying and preventing aging health risks in patients in schizophrenia.

## Acknowledgements/Funding

This work was supported by the Yale Physician Scientist Development Award (Z.M.H.) and CTSA (Z.M.H.; NIH UL1 TR001863), grants from the National Institute on Aging (A.H.C.; 1R01AG065403), the National Heart Lung Institute (J.L.S.; R01HL123915, R01HL155742, and 1R01HL169300), the National Institute of Diabetes and Digestive and Kidney Diseases (Z.M.H.; K23DK136932), and the Impetus Grants (R. Sehgal). Additionally, Dr. Harvanek is a Pepper Scholar with support from the Claude D. Pepper Older Americans Independence Center at Yale School of Medicine (P30AG021342). Dr. Higgins-Chen was supported by a Pilot Grant from the Claude D. Pepper Older Americans Independence Center at Yale School of Medicine (P30AG021342).

## Data availability

All data is available on NCBI GEO (https://www.ncbi.nlm.nih.gov/geo/) and datasets are listed in Table 1. Information on clozapine status can be found in their respective papers^13,60^.

## Code availability

Code to calculate all clocks except for OMICmAge and DNAmEMRAge are accessible at https://github.com/HigginsChenLab/methylCIPHER. Code to calculate OMICmAge, DNAmEMRAge and associated algorithms will be accessible via TruDiagnostic’s DNAm Analysis Software after publication. You can request access to the software at https://www.trudiagnostic.com/softwarerequest.

## Author contributions

Z.M.H. and A.H.C. designed the study and drafted the initial manuscript. Z.M.H. performed the analyses. R.S. contributed body plots and code for analyses. Z.M.H., A.H.C., R.S., D.B., J.K., and A.P. contributed to data cleaning and clock calculation. A.H.C. conceived the project and provided supervision. C.H.V. and M.P.B. provided data and conceptual input. M.J.C., V.B.D., J.L.S., and R.S. provided clock code and insights. All authors reviewed and contributed to the manuscript.

## References

1. Hennekens CH, Hennekens AR, Hollar D, Casey DE. Schizophrenia and increased risks of cardiovascular disease. Am Heart J. Dec 2005;150(6):1115–21. doi:10.1016/j.ahj.2005.02.007

2. Saha S, Chant D, McGrath J. A systematic review of mortality in schizophrenia: is the differential mortality gap worsening over time? Arch Gen Psychiatry. Oct 2007;64(10):1123–31. doi:10.1001/archpsyc.64.10.1123

3. Chan JKN, Correll CU, Wong CSM, et al. Life expectancy and years of potential life lost in people with mental disorders: a systematic review and meta-analysis. EClinicalMedicine. Nov 2023;65:102294. doi:10.1016/j.eclinm.2023.102294

4. Correll CU, Solmi M, Croatto G, et al. Mortality in people with schizophrenia: a systematic review and meta-analysis of relative risk and aggravating or attenuating factors. World Psychiatry. Jun 2022;21(2):248–271. doi:10.1002/wps.20994

5. Richmond-Rakerd LS, D’Souza S, Milne BJ, Caspi A, Moffitt TE. Longitudinal Associations of Mental Disorders With Physical Diseases and Mortality Among 2.3 Million New Zealand Citizens. JAMA Netw Open. Jan 4 2021;4(1):e2033448. doi:10.1001/jamanetworkopen.2020.33448

6. Halstead S, Cao C, Høgnason Mohr G, et al. Prevalence of multimorbidity in people with and without severe mental illness: a systematic review and meta-analysis. Lancet Psychiatry. Jun 2024;11(6):431–442. doi:10.1016/s2215-0366(24)00091-9

7. Pizzol D, Trott M, Butler L, et al. Relationship between severe mental illness and physical multimorbidity: a meta-analysis and call for action. BMJ Ment Health. Oct 2023;26(1)doi:10.1136/bmjment-2023-300870

8. Lu C, Jin D, Palmer N, et al. Large-scale real-world data analysis identifies comorbidity patterns in schizophrenia. Transl Psychiatry. Apr 11 2022;12(1):154. doi:10.1038/s41398-022-01916-y

9. Solmi M, Croatto G, Fornaro M, et al. Regional differences in mortality risk and in attenuating or aggravating factors in schizophrenia: A systematic review and meta-analysis. Eur Neuropsychopharmacol. Mar 2024;80:55–69. doi:10.1016/j.euroneuro.2023.12.010

10. Upthegrove R, Khandaker GM. Cytokines, Oxidative Stress and Cellular Markers of Inflammation in Schizophrenia. Curr Top Behav Neurosci. 2020;44:49–66. doi:10.1007/7854_2018_88

11. Hartwig FP, Borges MC, Horta BL, Bowden J, Davey Smith G. Inflammatory Biomarkers and Risk of Schizophrenia: A 2-Sample Mendelian Randomization Study. JAMA Psychiatry. Dec 1 2017;74(12):1226–1233. doi:10.1001/jamapsychiatry.2017.3191

12. Campeau A, Mills RH, Stevens T, et al. Multi-omics of human plasma reveals molecular features of dysregulated inflammation and accelerated aging in schizophrenia. Mol Psychiatry. Feb 2022;27(2):1217–1225. doi:10.1038/s41380-021-01339-z

13. Higgins-Chen AT, Boks MP, Vinkers CH, Kahn RS, Levine ME. Schizophrenia and Epigenetic Aging Biomarkers: Increased Mortality, Reduced Cancer Risk, and Unique Clozapine Effects. Biol Psychiatry. Aug 1 2020;88(3):224–235. doi:10.1016/j.biopsych.2020.01.025

14. Voisey J, Lawford BR, Morris CP, et al. Epigenetic analysis confirms no accelerated brain aging in schizophrenia. NPJ Schizophr. Sep 4 2017;3(1):26. doi:10.1038/s41537-017-0026-4

15. Oblak L, van der Zaag J, Higgins-Chen AT, Levine ME, Boks MP. A systematic review of biological, social and environmental factors associated with epigenetic clock acceleration. Ageing Res Rev. Aug 2021;69:101348. doi:10.1016/j.arr.2021.101348

16. Harvanek ZM, Boks MP, Vinkers CH, Higgins-Chen AT. The Cutting Edge of Epigenetic Clocks: In Search of Mechanisms Linking Aging and Mental Health. Biol Psychiatry. Nov 1 2023;94(9):694–705. doi:10.1016/j.biopsych.2023.02.001

17. McKinney BC, Lin H, Ding Y, Lewis DA, Sweet RA. DNA methylation age is not accelerated in brain or blood of subjects with schizophrenia. Schizophr Res. Jun 2018;196:39–44. doi:10.1016/j.schres.2017.09.025

18. Caspi A, Shireby G, Mill J, Moffitt TE, Sugden K, Hannon E. Accelerated Pace of Aging in Schizophrenia: Five Case-Control Studies. Biol Psychiatry. Jun 1 2024;95(11):1038–1047. doi:10.1016/j.biopsych.2023.10.023

19. Sluiskes MH, Goeman JJ, Beekman M, Slagboom PE, Putter H, Rodríguez-Girondo M. Clarifying the biological and statistical assumptions of cross-sectional biological age predictors: an elaborate illustration using synthetic and real data. BMC Med Res Methodol. Mar 8 2024;24(1):58. doi:10.1186/s12874-024-02181-x

20. Lu AT, Quach A, Wilson JG, et al. DNA methylation GrimAge strongly predicts lifespan and healthspan. Aging (Albany NY). Jan 21 2019;11(2):303–327. doi:10.18632/aging.101684

21. Nelson PG, Promislow DEL, Masel J. Biomarkers for Aging Identified in Cross-sectional Studies Tend to Be Non-causative. J Gerontol A Biol Sci Med Sci. Feb 14 2020;75(3):466–472. doi:10.1093/gerona/glz174

22. Schaie KW. Age changes and age differences. Gerontologist. Jun 1967;7(2):128-32. doi:10.1093/geront/7.2_part_1.128

23. Ying K, Liu H, Tarkhov AE, et al. Causality-enriched epigenetic age uncouples damage and adaptation. Nat Aging. Feb 2024;4(2):231–246. doi:10.1038/s43587-023-00557-0

24. Chen Q, Dwaraka VB, Carreras-Gallo N, et al. OMICmAge: An integrative multi-omics approach to quantify biological age with electronic medical records. bioRxiv. 2023:2023.10.16.562114. doi:10.1101/2023.10.16.562114

25. Tomusiak A, Floro A, Tiwari R, et al. Development of an epigenetic clock resistant to changes in immune cell composition. Commun Biol. Aug 2 2024;7(1):934. doi:10.1038/s42003-024-06609-4

26. Ndhlovu LC, Bendall ML, Dwaraka V, et al. Retro-age: A unique epigenetic biomarker of aging captured by DNA methylation states of retroelements. Aging Cell. Aug 2 2024:e14288. doi:10.1111/acel.14288

27. Boks MP, Houtepen LC, Xu Z, et al. Genetic vulnerability to DUSP22 promoter hypermethylation is involved in the relation between in utero famine exposure and schizophrenia. NPJ Schizophr. Aug 21 2018;4(1):16. doi:10.1038/s41537-018-0058-4

28. Hannon E, Dempster EL, Mansell G, et al. DNA methylation meta-analysis reveals cellular alterations in psychosis and markers of treatment-resistant schizophrenia. Elife. Feb 26 2021;10doi:10.7554/eLife.58430

29. Thrush KL, Higgins-Chen AT, Liu Z, Levine ME. R methylCIPHER: A Methylation Clock Investigational Package for Hypothesis-Driven Evaluation & Research. bioRxiv. 2022:2022.07.13.499978. doi:10.1101/2022.07.13.499978

30. Higgins-Chen AT, Thrush KL, Wang Y, et al. A computational solution for bolstering reliability of epigenetic clocks: implications for clinical trials and longitudinal tracking. Nature Aging. 2022/07/01 2022;2(7):644-661. doi:10.1038/s43587-022-00248-2

31. Sehgal R, Markov Y, Qin C, et al. Systems Age: a single blood methylation test to quantify aging heterogeneity across 11 physiological systems. Nat Aging. Sep 2025;5(9):1880–1896. doi:10.1038/s43587-025-00958-3

32. Chen Q, Dwaraka VB, Carreras-Gallo N, et al. OMICmAge: An integrative multi-omics approach to quantify biological age with electronic medical records. bioRxiv. Oct 24 2023;doi:10.1101/2023.10.16.562114

33. Teschendorff AE, Breeze CE, Zheng SC, Beck S. A comparison of reference-based algorithms for correcting cell-type heterogeneity in Epigenome-Wide Association Studies. BMC Bioinformatics. Feb 13 2017;18(1):105. doi:10.1186/s12859-017-1511-5

34. R: A Language and Environment for Statistical Computing. R Foundation for Statistical Computing; 2020. https://www.R-project.org/

35. Viechtbauer W. Conducting Meta-Analyses in R with the metafor Package. Journal of Statistical Software. 08/05 2010;36(3):1 - 48. doi:10.18637/jss.v036.i03

36. Wickham H. ggplot2 : Elegant Graphics for Data Analysis. 2nd ed. Springer International Publishing : Imprint: Springer,; 2016:XVI, 260 pages 232 illustrations, 140 illustrations in color.

37. Chen BH, Marioni RE, Colicino E, et al. DNA methylation-based measures of biological age: meta-analysis predicting time to death. Aging (Albany NY). Sep 28 2016;8(9):1844–1865. doi:10.18632/aging.101020

38. Tian YE, Di Biase MA, Mosley PE, et al. Evaluation of Brain-Body Health in Individuals With Common Neuropsychiatric Disorders. JAMA Psychiatry. Jun 1 2023;80(6):567–576. doi:10.1001/jamapsychiatry.2023.0791

39. Rodrigues M, Wiener JC, Stranges S, Ryan BL, Anderson KK. The risk of physical multimorbidity in people with psychotic disorders: A systematic review and meta-analysis. J Psychosom Res. Jan 2021;140:110315. doi:10.1016/j.jpsychores.2020.110315

40. de Leon J, Diaz FJ. A meta-analysis of worldwide studies demonstrates an association between schizophrenia and tobacco smoking behaviors. Schizophr Res. Jul 15 2005;76(2-3):135–57. doi:10.1016/j.schres.2005.02.010

41. Chow RTS, Whiting D, Favril L, Ostinelli E, Cipriani A, Fazel S. An umbrella review of adverse effects associated with antipsychotic medications: the need for complementary study designs. Neurosci Biobehav Rev. Dec 2023;155:105454. doi:10.1016/j.neubiorev.2023.105454

42. Gillespie AL, Walker EM, Hannon E, et al. Longitudinal changes in DNA methylation associated with clozapine use in treatment-resistant schizophrenia from two international cohorts. Transl Psychiatry. Sep 27 2024;14(1):390. doi:10.1038/s41398-024-03102-8

43. Luykx JJ, Gonzalez-Diaz JM, Guu TW, et al. An international research agenda for clozapine-resistant schizophrenia. Lancet Psychiatry. Aug 2023;10(8):644–652. doi:10.1016/s2215-0366(23)00109-8

44. Conole ELS, Stevenson AJ, Muñoz Maniega S, et al. DNA Methylation and Protein Markers of Chronic Inflammation and Their Associations With Brain and Cognitive Aging. Neurology. Dec 7 2021;97(23):e2340–e2352. doi:10.1212/wnl.0000000000012997

45. Jiang Y, Cai Y, Lu Y, Wu G, Zhang XY. Relationship between anemia and its correlates and cognitive function in Chinese patients with chronic schizophrenia: A large cross-sectional study. Schizophr Res Cogn. Jun 2024;36:100300. doi:10.1016/j.scog.2024.100300

46. Xu H, Zheng L, Wang L, Gao H, Wei Y, Chen J. Albumin and Associated Biomarkers in Severe Neuropsychiatric Disorders: Acute-Phase Schizophrenia and Bipolar Disorder. Neuropsychiatr Dis Treat. 2023;19:2027–2037. doi:10.2147/ndt.S423399

47. Hao M, Jiang S, Tang J, et al. Ratio of Red Blood Cell Distribution Width to Albumin Level and Risk of Mortality. JAMA Netw Open. May 1 2024;7(5):e2413213. doi:10.1001/jamanetworkopen.2024.13213

48. Khan S. IGFBP-2 Signaling in the Brain: From Brain Development to Higher Order Brain Functions. Front Endocrinol (Lausanne). 2019;10:822. doi:10.3389/fendo.2019.00822

49. Nolin MA, Demers MF, Rauzier C, et al. Circulating IGFBP-2 levels reveal atherogenic metabolic risk in schizophrenic patients using atypical antipsychotics. World J Biol Psychiatry. Mar 2021;22(3):175–182. doi:10.1080/15622975.2020.1770858

50. Zong X, Fan Q, Yang Q, Pan R, Zhuang L, Tao R. Phenylacetylglutamine as a risk factor and prognostic indicator of heart failure. ESC Heart Fail. Aug 2022;9(4):2645–2653. doi:10.1002/ehf2.13989

51. Liu G, Nguyen NQH, Wong KE, et al. Metabolomic Association and Risk Prediction With Heart Failure in Older Adults. Circ Heart Fail. Mar 2024;17(3):e010896. doi:10.1161/circheartfailure.123.010896

52. Claesen K, Mertens JC, Leenaerts D, Hendriks D. Carboxypeptidase U (CPU, TAFIa, CPB2) in Thromboembolic Disease: What Do We Know Three Decades after Its Discovery? Int J Mol Sci. Jan 17 2021;22(2)doi:10.3390/ijms22020883

53. Contrepois K, Mann C, Fenaille F. H2B Type 1-K Accumulates in Senescent Fibroblasts with Persistent DNA Damage along with Methylated and Phosphorylated Forms of HMGA1. Proteomes. Jun 21 2021;9(2)doi:10.3390/proteomes9020030

54. Cao HM, Ye XP, Ma JH, et al. Mimecan, a Hormone Abundantly Expressed in Adipose Tissue, Reduced Food Intake Independently of Leptin Signaling. EBioMedicine. Nov 2015;2(11):1718–24. doi:10.1016/j.ebiom.2015.09.044

55. Oh HS, Rutledge J, Nachun D, et al. Organ aging signatures in the plasma proteome track health and disease. Nature. Dec 2023;624(7990):164–172. doi:10.1038/s41586-023-06802-1

56. Ahn J, Suh Y, Lee K. Chordin-like 1, a Novel Adipokine, Markedly Promotes Adipogenesis and Lipid Accumulation. Cells. Feb 15 2023;12(4)doi:10.3390/cells12040624

57. Yang J, Chen T, Sun L, et al. Potential metabolite markers of schizophrenia. Mol Psychiatry. Jan 2013;18(1):67–78. doi:10.1038/mp.2011.131

58. Li Y, Ma C, Li S, et al. Regulatory Variant rs2535629 in ITIH3 Intron Confers Schizophrenia Risk By Regulating CTCF Binding and SFMBT1 Expression. Adv Sci (Weinh). Feb 2022;9(6):e2104786. doi:10.1002/advs.202104786

59. Li K, Li Y, Wang J, et al. A functional missense variant in ITIH3 affects protein expression and neurodevelopment and confers schizophrenia risk in the Han Chinese population. J Genet Genomics. May 20 2020;47(5):233–248. doi:10.1016/j.jgg.2020.04.001

60. Gunasekara CJ, Hannon E, MacKay H, et al. A machine learning case-control classifier for schizophrenia based on DNA methylation in blood. Transl Psychiatry. Aug 3 2021;11(1):412. doi:10.1038/s41398-021-01496-3

