## Supplementary Materials for "Multidimensional Epigenetic Clocks Reveal Physiological System-Specific Aging in Schizophrenia"

### Supplementary Methods:

#### Clock Calculation

Causality-enriched clocks were calculated as described by Ying et al, 2024^1^. Systems-based clocks were calculated as described in Sehgal et al, 2024^2^. OMICmAge, OMICmAge epigenetic biomarker proxies (EBPs), DNAmEMRAge, RetroelementAge, IntrinClock, and other clocks were calculated using code provided by the authors^3,4^. All scores were incorporated into the methylCIPHER package (github.com/HigginsChenLab/methylCIPHER) for standardized implementation^5^. The methylCIPHER package was also used to calculate earlier iterations of clocks, including PC clocks^6^. Of note, OMICmAge, DNAmEMRAge, and RetroelementAge were trained on EPICv1 data. Although many CpGs were missing (OMICmAge 45% missing, EBPs 34-57% missing (mean 50%), DNAmEMRAge 50% missing, RetroelementAge 70% missing), it is known that much of the aging signal is preserved on 450K data (as assessed by correlation between clocks calculated using full EPIC data versus clocks calculated using only 450K CpGs, and by preserved prediction of aging outcomes)^3^. For RetroelementAge, calculations were adjusted based on the array used in the dataset as suggested by the authors. See supplementary table 1 for details on all Clocks used.

### Supplementary Figure Legends:

#### Supplementary Figure 1: Distribution of ages across studies

Histograms of ages of both controls (red) and patients with schizophrenia-spectrum disorders (blue) by study.

#### Supplementary Figure 2: Meta-analysis results of the effect of schizophrenia on epigenetic clocks and DNAm proxies

Forest plots of meta-analysis results all clocks, including novel clocks, traditional clocks, and DNAm Proxies, with positive numbers indicating epigenetic age is higher in patients with schizophrenia compared to controls. Nominal 95^th^ percentile confidence intervals are presented.

#### Supplementary Figure 3: Heatmap summary of relationship between traditional clocks and schizophrenia

accounting for 14 traditional clocks, nominal unadjusted p of 0.00357 = Bonferroni adjusted p of 0.05.

#### Supplementary Figure 4: Meta-analysis results of the effect of clozapine treatment on epigenetic clocks and DNAm proxies

Forest plots of meta-analysis results all clocks, including novel clocks, traditional clocks, and DNAm Proxies, with positive numbers indicating epigenetic age is higher in patients with schizophrenia on Clozapine compared to patients with schizophrenia not on clozapine. Nominal 95^th^ percentile confidence intervals are presented.

1.	Ying K, Liu H, Tarkhov AE, et al. Causality-enriched epigenetic age uncouples damage and adaptation. *Nat Aging.* 2024;4(2):231-246.

2.	Sehgal R, Markov Y, Qin C, et al. Systems Age: A single blood methylation test to quantify aging heterogeneity across 11 physiological systems. *bioRxiv.* 2024:2023.2007.2013.548904.

3.	Chen Q, Dwaraka VB, Carreras-Gallo N, et al. OMICmAge: An integrative multi-omics approach to quantify biological age with electronic medical records. *bioRxiv.* 2023.

4.	Ndhlovu LC, Bendall ML, Dwaraka V, et al. Retroelement-Age Clocks: Epigenetic Age Captured by Human Endogenous Retrovirus and LINE-1 DNA methylation states. *bioRxiv.* 2023.

5.	Thrush KL, Higgins-Chen AT, Liu Z, Levine ME. R methylCIPHER: A Methylation Clock Investigational Package for Hypothesis-Driven Evaluation &amp; Research. *bioRxiv.* 2022:2022.2007.2013.499978.

6.	Higgins-Chen AT, Thrush KL, Wang Y, et al. A computational solution for bolstering reliability of epigenetic clocks: implications for clinical trials and longitudinal tracking. *Nature Aging.* 2022;2(7):644-661.
