## Supplementary figures and images for "Multidimensional Epigenetic Clocks Reveal Physiological System-Specific Aging in Schizophrenia"

### Supplementary Figure 1

#### Supplementary Figure 1

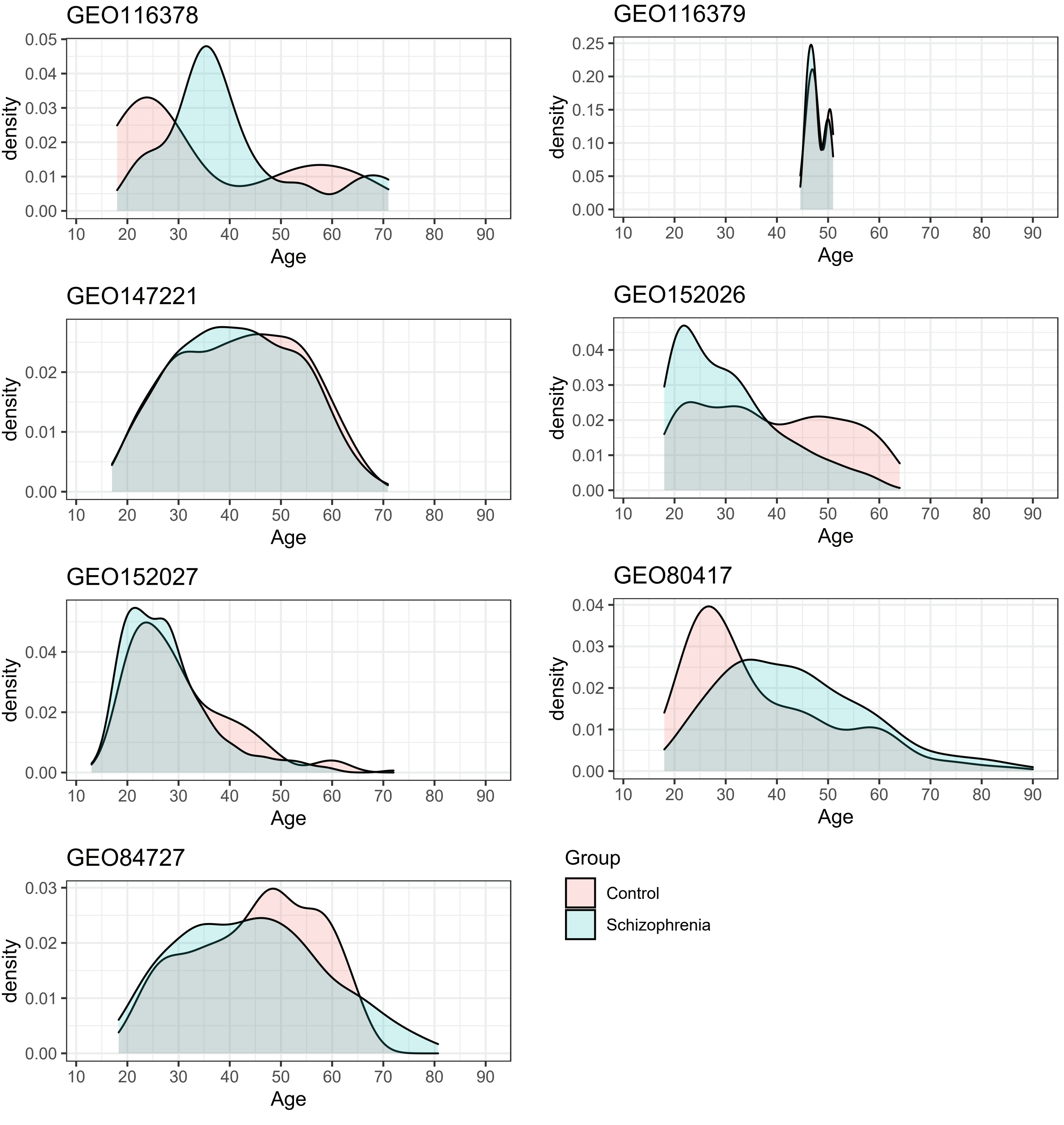

### Supplementary Figure 2

#### Supplementary Figure 2

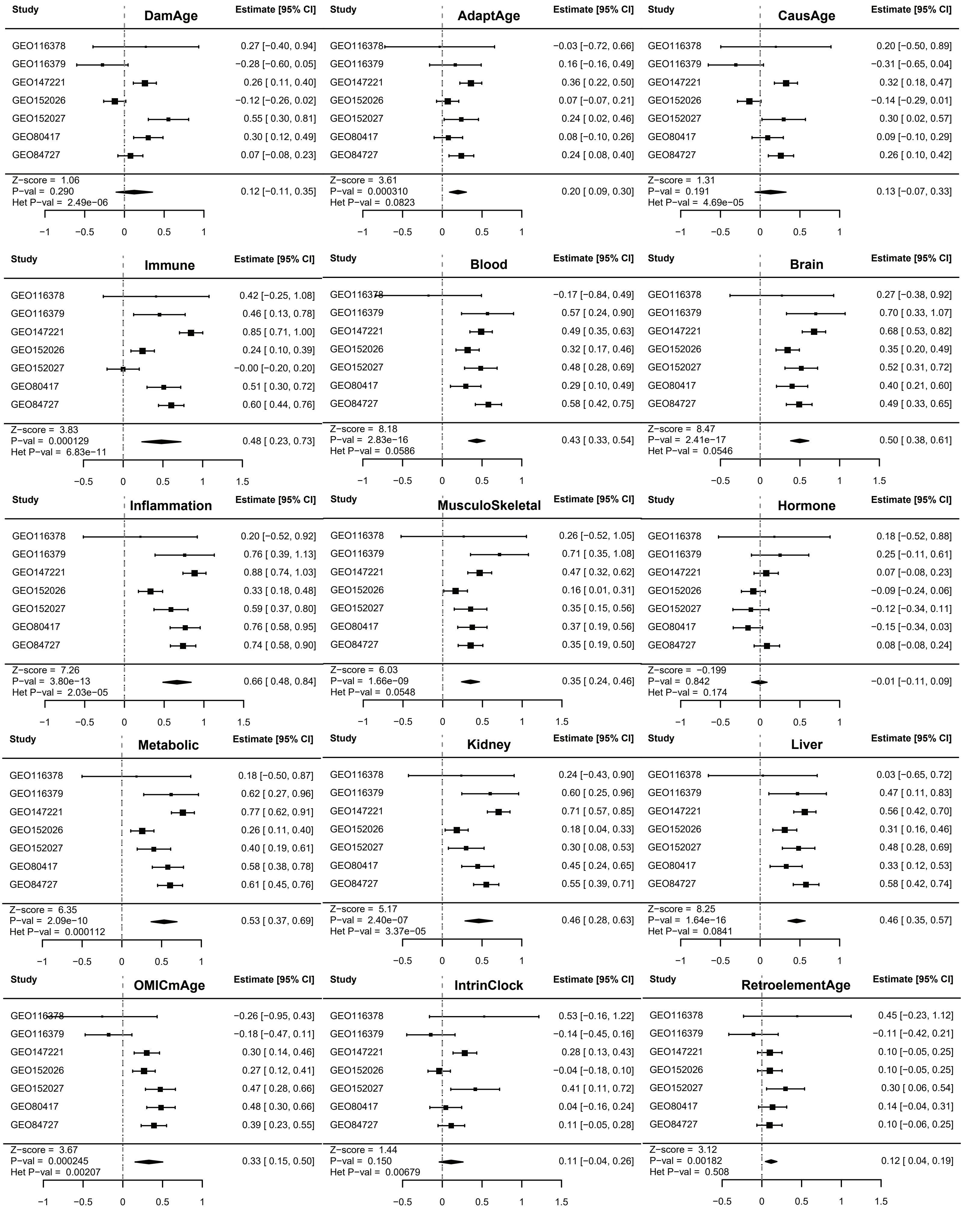

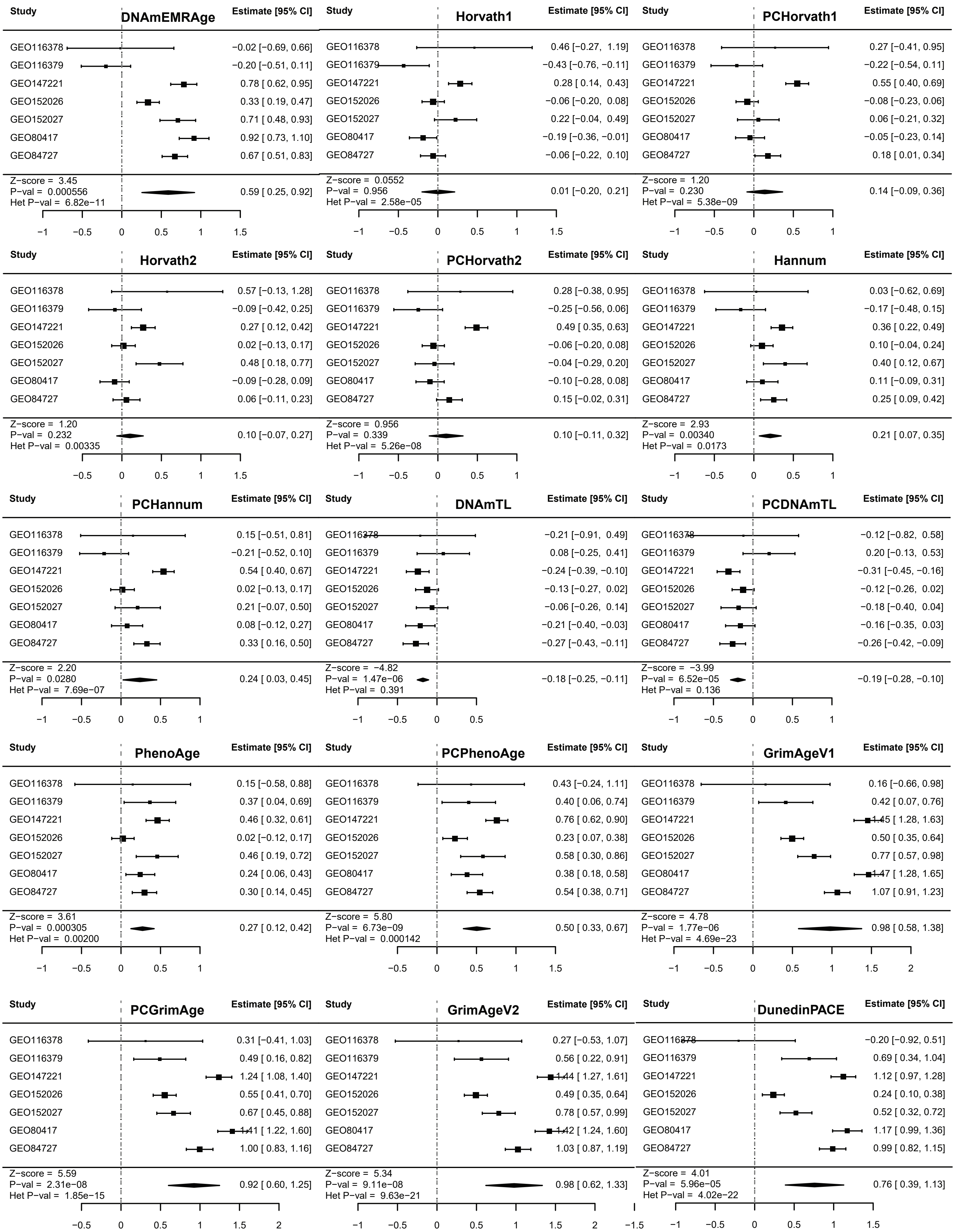

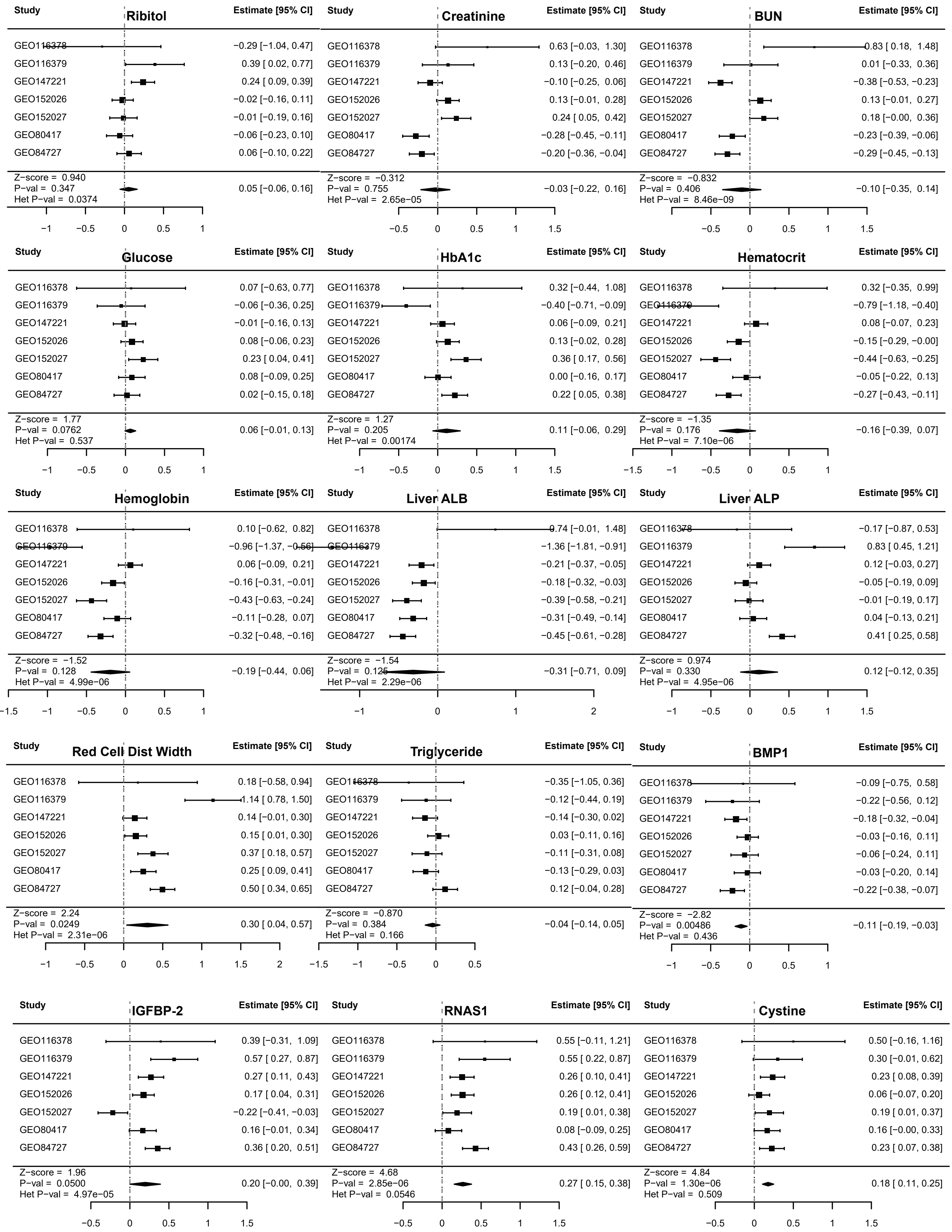

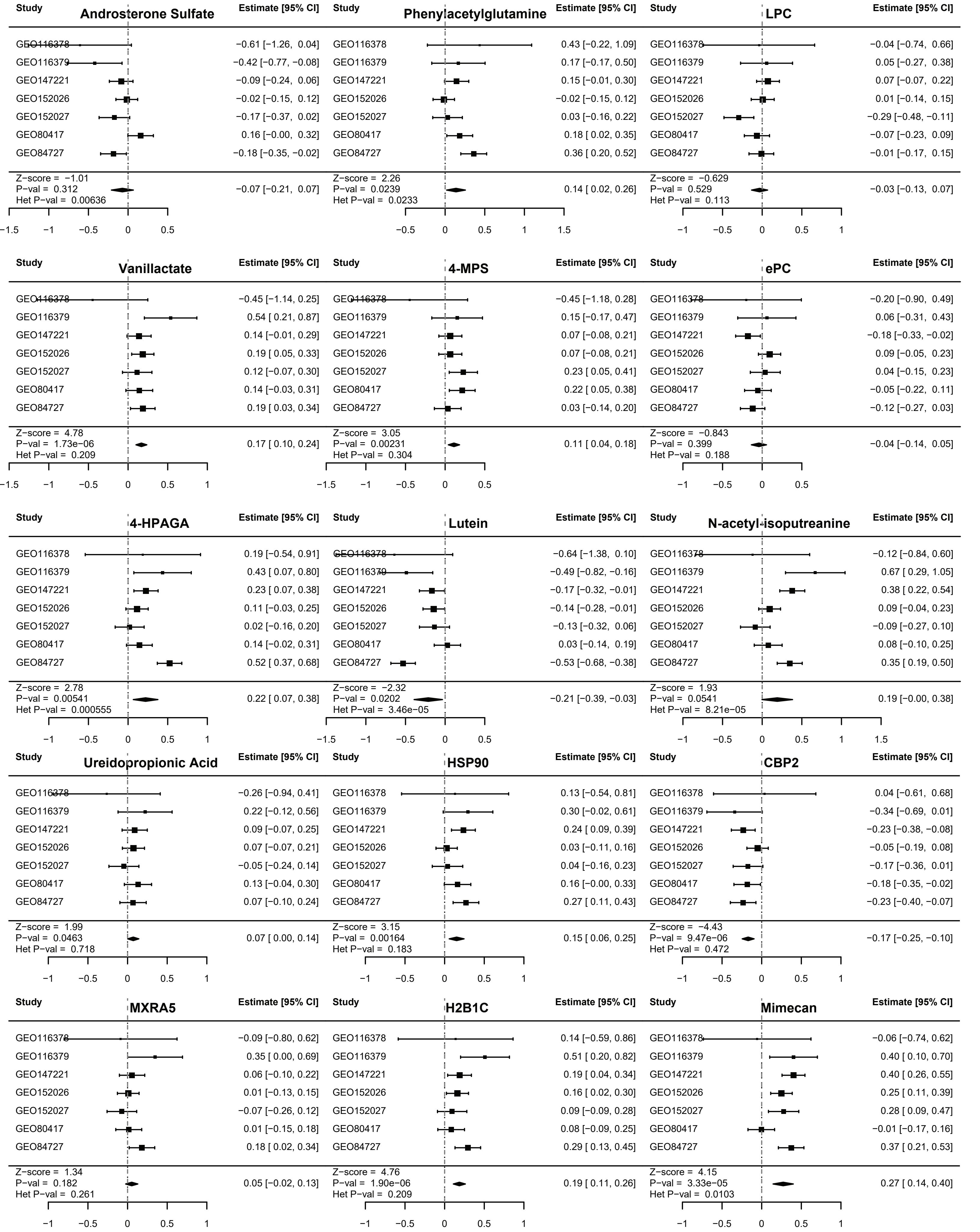

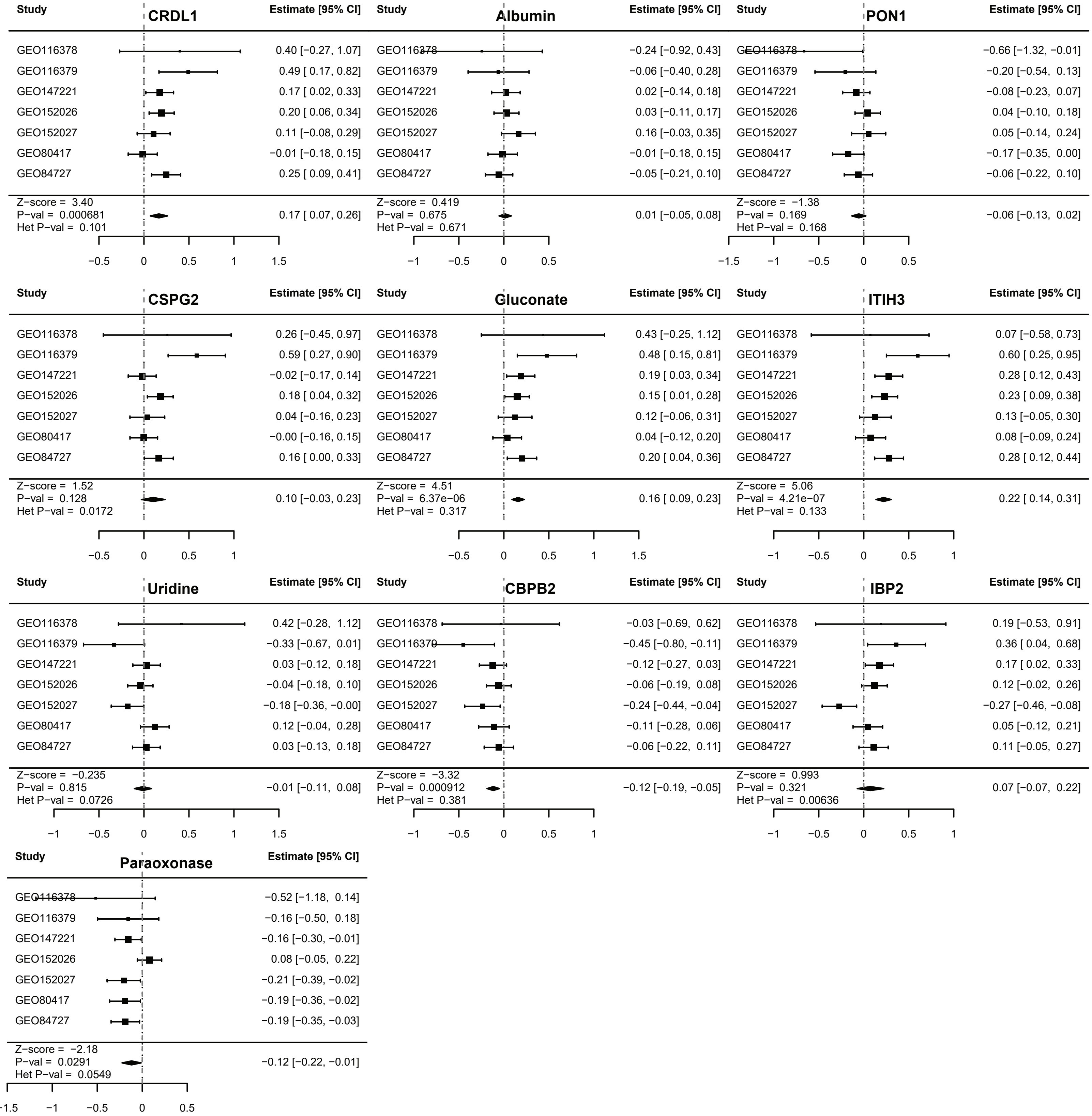

### Supplementary Figure 3

**Supplementary Figure 3**

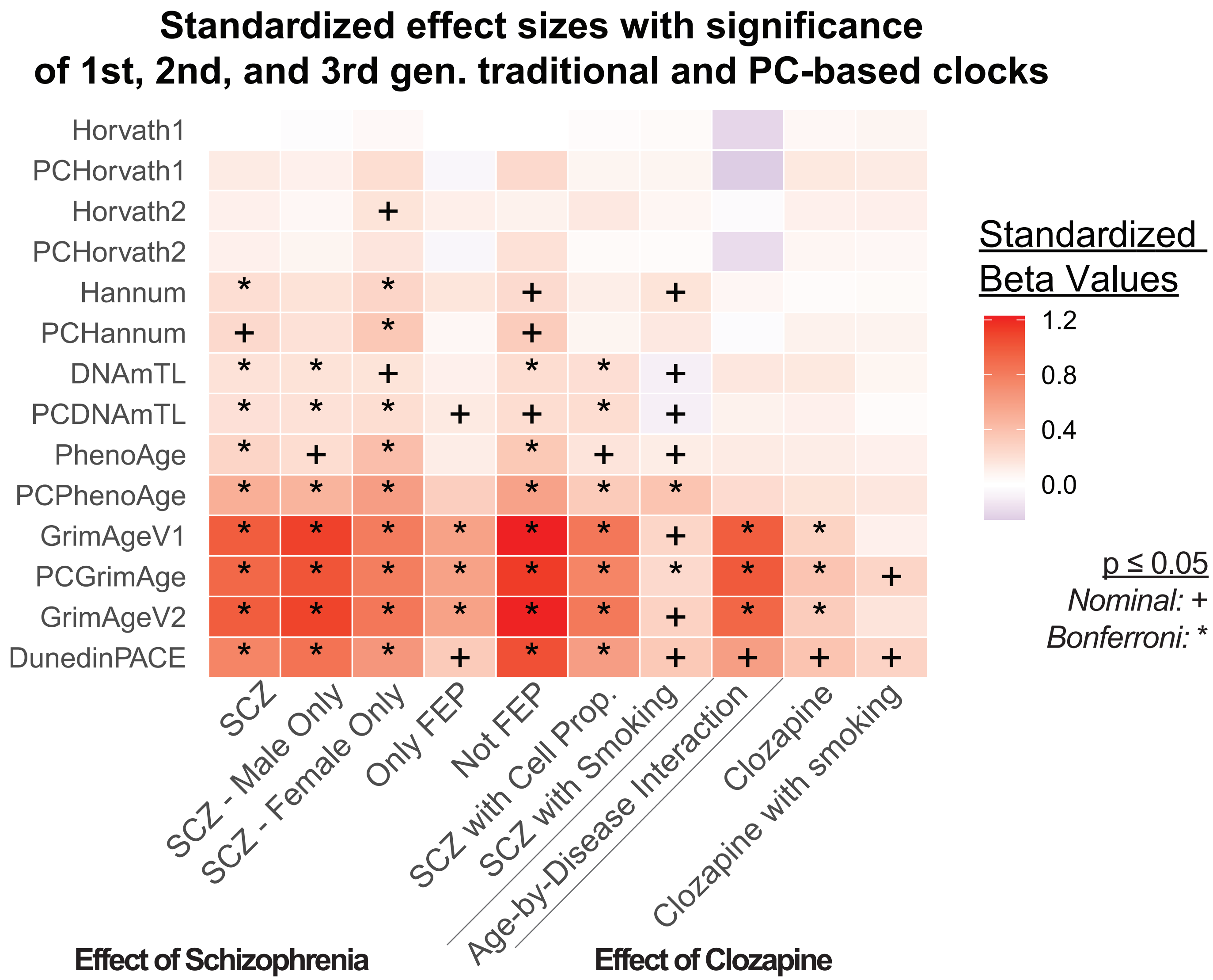

### Supplementary Figure 4

#### Supplementary Figure 4

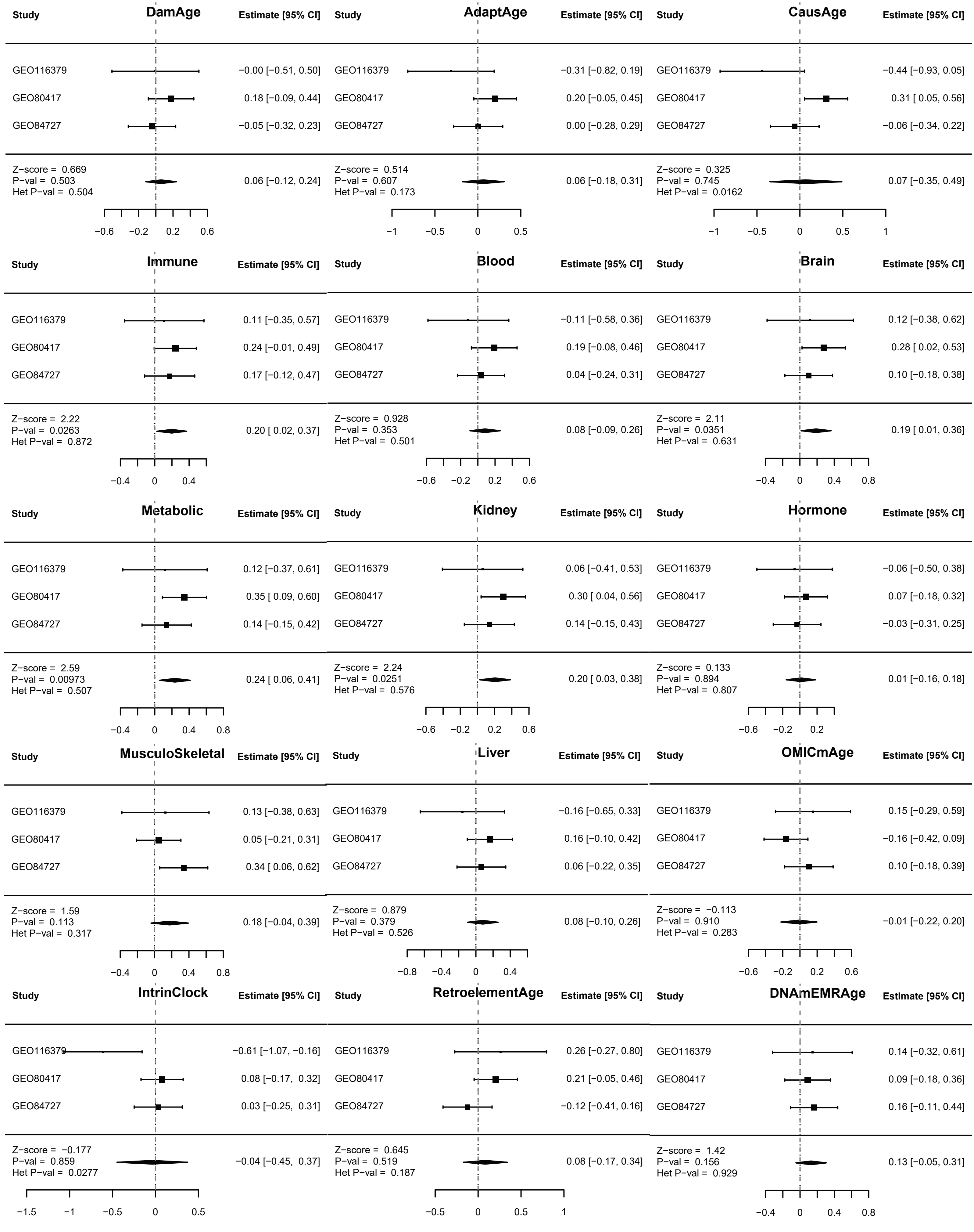

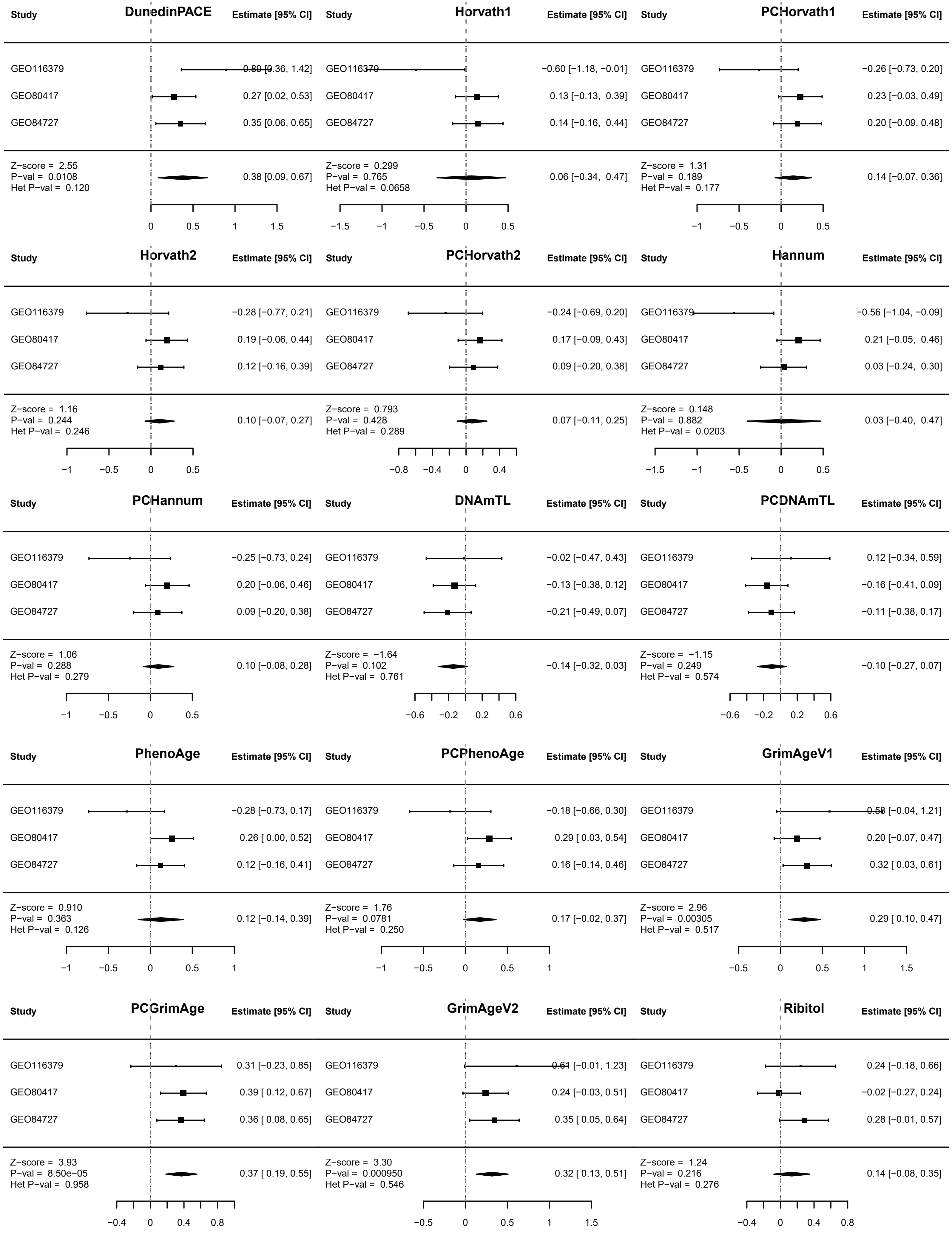

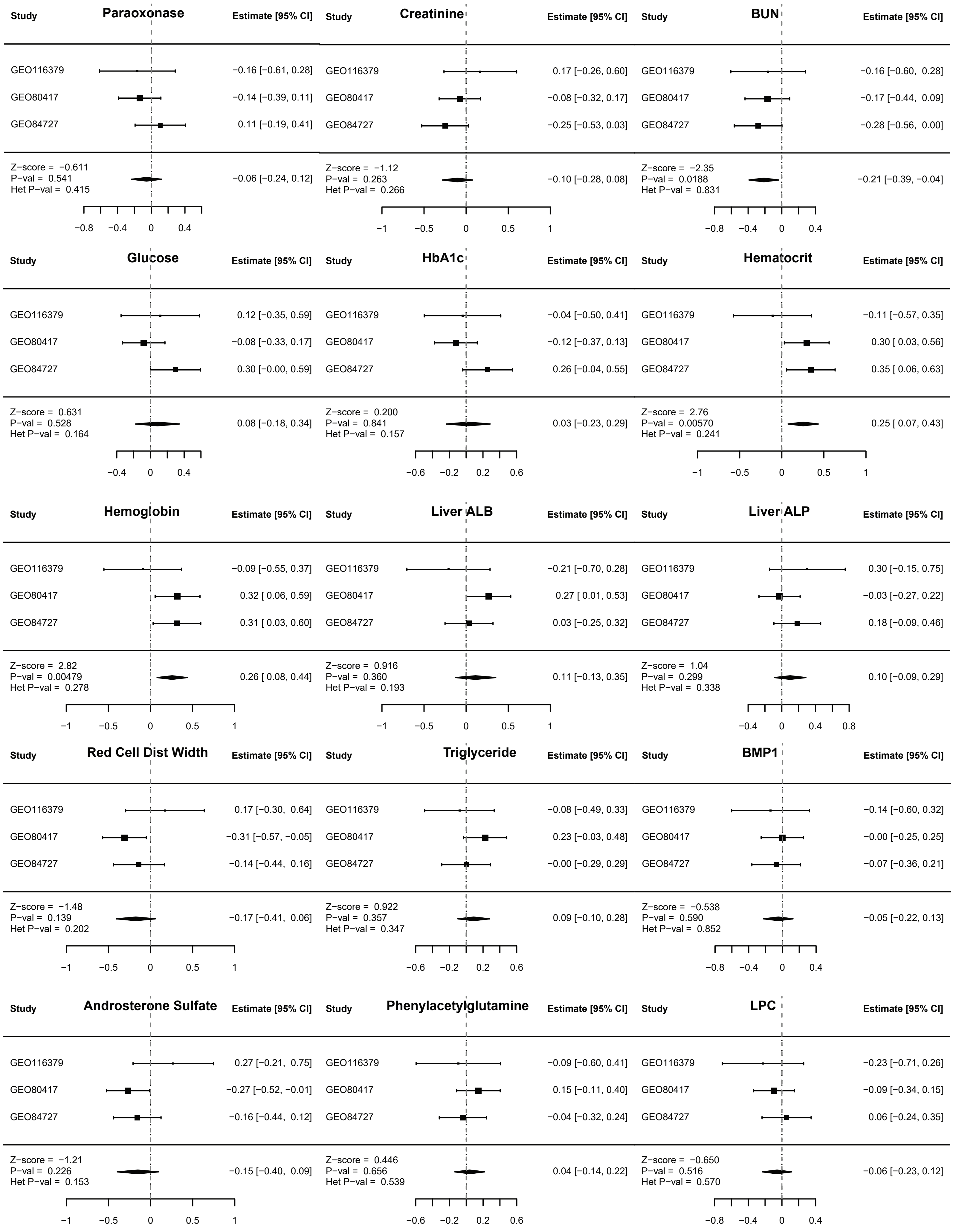

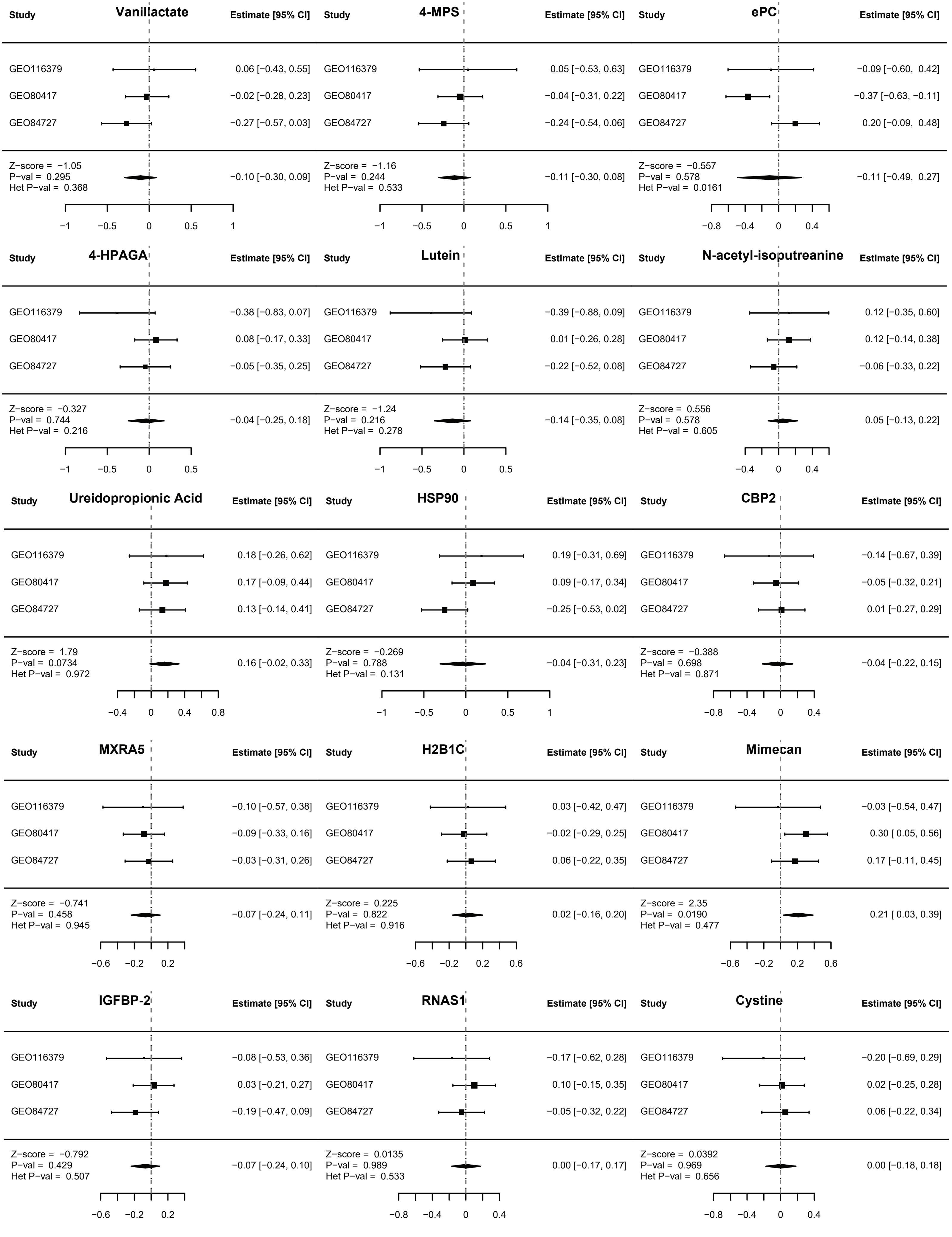

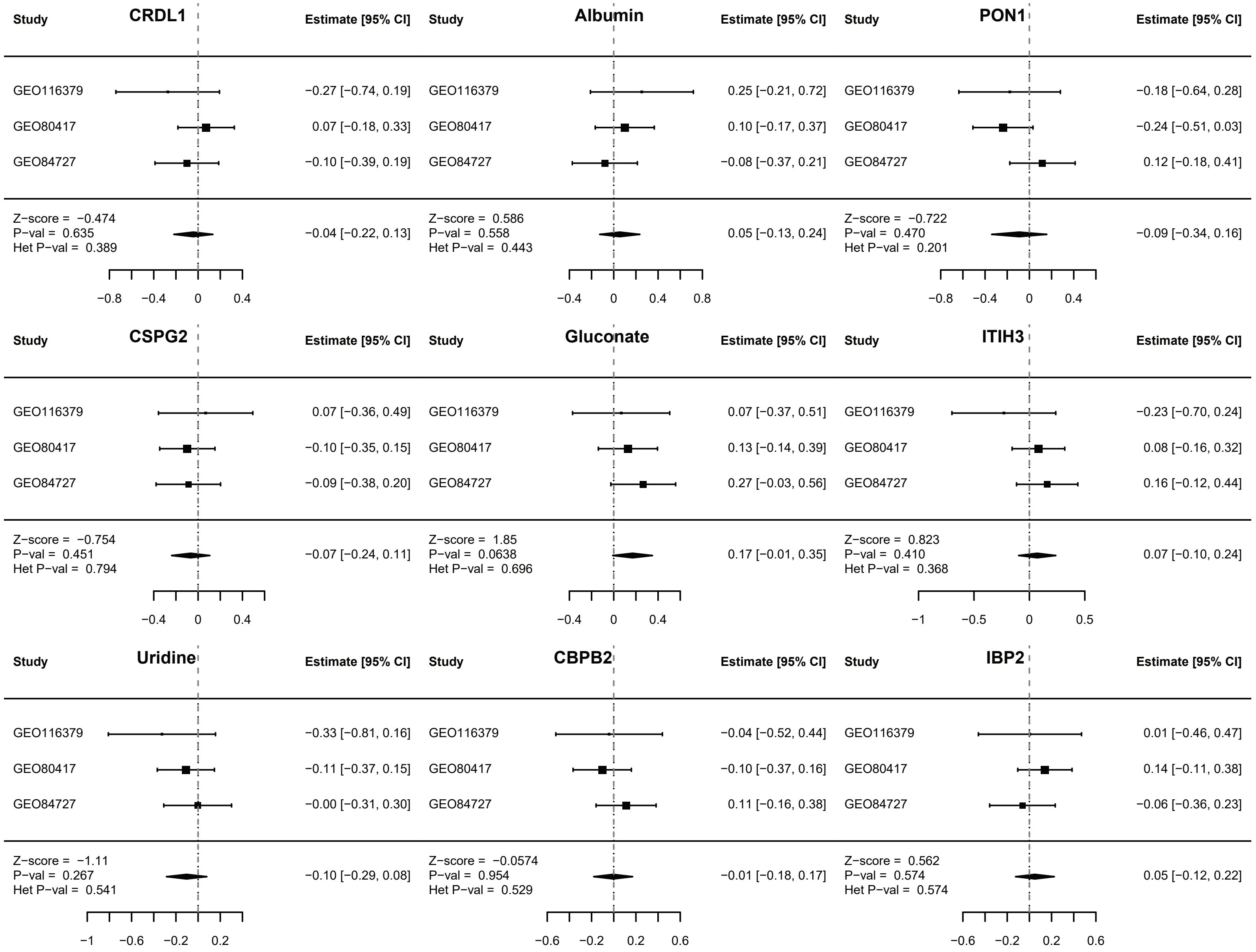
